# Innate lymphoid cells and disease tolerance in SARS-CoV-2 infection

**DOI:** 10.1101/2021.01.14.21249839

**Authors:** Noah J. Silverstein, Yetao Wang, Zachary Manickas-Hill, Claudia Carbone, Ann Dauphin, Brittany P. Boribong, Maggie Loiselle, Jameson Davis, Maureen M. Leonard, Leticia Kuri-Cervantes, MGH COVID-19 Collection & Processing Team, Nuala J. Meyer, Michael R. Betts, Jonathan Z. Li, Bruce Walker, Xu G. Yu, Lael M. Yonker, Jeremy Luban

## Abstract

Risk of severe COVID-19 increases with age, is greater in males, and is associated with lymphopenia, but not with higher burden of SARS-CoV-2. It is unknown whether effects of age and sex on abundance of specific lymphoid subsets explain these correlations. This study found that the abundance of innate lymphoid cells (ILCs) decreases more than 7-fold over the human lifespan — T cell subsets decrease less than 2-fold — and is lower in males than in females. After accounting for effects of age and sex, ILCs, but not T cells, were lower in adults hospitalized with COVID-19, independent of lymphopenia. Among SARS-CoV-2-infected adults, the abundance of ILCs, but not of T cells, correlated inversely with odds and duration of hospitalization, and with severity of inflammation. ILCs were also uniquely decreased in pediatric COVID-19 and the numbers of these cells did not recover during follow-up. In contrast, children with MIS-C had depletion of both ILCs and T cells, and both cell types increased during follow-up. In both pediatric COVID-19 and MIS-C, ILC abundance correlated inversely with inflammation. Blood ILC mRNA and phenotype tracked closely with ILCs from lung. Importantly, blood ILCs produced amphiregulin, a protein implicated in disease tolerance and tissue homeostasis, and the percentage of amphiregulin-producing ILCs was higher in females than in males. These results suggest that, by promoting disease tolerance, homeostatic ILCs decrease morbidity and mortality associated with SARS-CoV-2 infection, and that lower ILC abundance accounts for increased COVID-19 severity with age and in males.

## INTRODUCTION

The risk of severe COVID-19 and death in people infected with SARS-CoV-2 increases with age and is greater in men than in women (Alkhouli et al., 2020; Bunders and Altfeld, 2020; Gupta et al., 2021; Laxminarayan et al., 2020; Mauvais-Jarvis, 2020; O’Driscoll et al., 2020; Peckham et al., 2020; Richardson et al., 2020; Scully et al., 2020). These trends have been observed in people infected with SARS-CoV (Chen and Subbarao, 2007; Donnelly et al., 2003; Karlberg, 2004), or with MERS-CoV (Alghamdi et al., 2014), and in laboratory animals challenged with SARS-CoV or SARS-CoV-2 (Channappanavar et al., 2017; Leist et al., 2020). Yet, the mechanisms underlying these effects of age and sex on COVID-19 morbidity and mortality remain poorly understood.

The composition and function of the human immune system changes with age and exhibits sexual dimorphism (Darboe et al., 2020; Klein and Flanagan, 2016; Márquez et al., 2020; Patin et al., 2018; Solana et al., 2012), with consequences for survival of infection, response to vaccination, and susceptibility to autoimmune disease (Flanagan et al., 2017; Giefing-Kröll et al., 2015; Márquez et al., 2020; Mauvais-Jarvis, 2020; Patin et al., 2018; Piasecka et al., 2018). Better understanding of these effects might provide clues as to why the clinical outcome of SARS-CoV-2 infection is so variable, ranging from asymptomatic to lethal (Cevik et al., 2021; He et al., 2021; Jones et al., 2021; Lee et al., 2020; Lennon et al., 2020; Ra et al., 2021; Richardson et al., 2020; Yang et al., 2021).

Survival after infection with a pathogenic virus such as SARS-CoV-2 requires not only that the immune system control and eliminate the pathogen, but that disease tolerance mechanisms limit tissue damage caused by the pathogen or by host inflammatory responses (Ayres, 2020a; McCarville and Ayres, 2018; Medzhitov et al., 2012; Schneider and Ayres, 2008). Research with animal models has demonstrated that genetic and environmental factors can promote host fitness without directly inhibiting pathogen replication (Ayres, 2020a; Cumnock et al., 2018; Jhaveri et al., 2007; McCarville and Ayres, 2018; Medzhitov et al., 2012; Råberg et al., 2007; Sanchez et al., 2018; Schneider and Ayres, 2008; Wang et al., 2016). Although in most cases the underlying mechanism is unknown, some of these models suggest that subsets of innate lymphoid cells (ILCs) contribute to disease tolerance (Artis and Spits, 2015; Branzk et al., 2018; Califano et al., 2018; Diefenbach et al., 2020; McCarville and Ayres, 2018; Monticelli et al., 2015, 2011).

ILCs lack clonotypic antigen receptors but overlap developmentally and functionally with T cells. Based on expression of characteristic transcription factors and specific inducible cytokines, ILCs are classified into ILC1, ILC2, and ILC3 subsets that are analogous to T_H_1, T_H_2, and T_H_17 cells respectively (Artis and Spits, 2015; Cherrier et al., 2018; Vivier et al., 2018; Yudanin et al., 2019). Additionally, some ILC subsets produce the epidermal growth factor family member amphiregulin (AREG) that maintains the integrity of epithelial barriers in the lung and intestine (Branzk et al., 2018; Jamieson et al., 2013; Monticelli et al., 2015, 2011), and promotes tissue repair (Artis and Spits, 2015; Cherrier et al., 2018; Klose and Artis, 2016; Rak et al., 2016). In models of influenza infection in mice, homeostatic ILCs and exogenous AREG promote lung epithelial integrity, decrease disease severity, and increase survival, without decreasing pathogen burden (Califano et al., 2018; Jamieson et al., 2013; Monticelli et al., 2011).

Little is known about disease tolerance in the context of human infectious diseases. Interestingly, SARS-CoV-2 viral load does not reliably discriminate symptomatic from asymptomatic infection (Cevik et al., 2021; Jones et al., 2021; Lee et al., 2020; Lennon et al., 2021; Ra et al., 2021; Yang et al., 2021). This discrepancy between SARS-CoV-2 viral load and the severity of COVID-19 is especially pronounced in children, who rarely have severe COVID-19 (Charles Bailey et al., 2020; Li et al., 2020; Lu et al., 2020; Poline et al., 2020), though viral load may be comparable to that in adults with severe COVID-19 (Heald-Sargent et al., 2020; LoTempio et al., 2021; Yonker et al., 2020). These observations suggest that age-dependent, disease tolerance mechanisms influence the severity of COVID-19. In mice, homeostatic ILCs decrease in abundance in the lung with increasing age, and lose their ability to maintain disease tolerance during influenza infection (D’Souza et al., 2019). Although the distribution of ILCs within human tissues differs from mice and is heterogeneous among individuals (Yudanin et al., 2019), human ILCs share many features with those in mice (Vivier et al., 2018) and therefore likely perform similar roles in maintaining tissue homeostasis and disease tolerance.

ILCs in peripheral blood have been reported to be depleted in individuals with severe COVID-19 (García et al., 2020; Kuri-Cervantes et al., 2020), but it is difficult to determine the extent to which ILCs are decreased independently from the overall lymphopenia associated with COVID-19 (Chen et al., 2020; Huang et al., 2020; Huang and Pranata, 2020; Zhang et al., 2020; Zhao et al., 2020), or from the reported decreases in other blood lymphoid cell populations (Kuri-Cervantes et al., 2020; Lucas et al., 2020; Mathew et al., 2020; Mudd et al., 2020; Zheng et al., 2020). In addition, assessment of lymphoid cell abundance, in the context of a disease for which age and sex are risk factors for severity, is confounded by programmed differences in lymphocyte abundance with age and sex (Márquez et al., 2020; Patin et al., 2018). The goal of this study was to determine whether the abundance of any blood lymphoid cell population was altered in COVID-19, independent of age, sex, and global lymphopenia, and whether abundance of any lymphoid cell population correlated with clinical outcome in SARS-CoV-2 infection.

## RESULTS

### Characteristics of adult blood donors hospitalized for COVID-19, treated for COVID-19 as outpatients, or SARS-CoV-2-uninfected controls

The first group of blood donors in this study included SARS-CoV-2-infected adults hospitalized for severe COVID-19 (N = 40), among whom 33 (82.5%) were admitted to the ICU, 32 (80%) required intubation with mechanical ventilation, and 7 (17.5%) died (Table 1). This group had a mean age of 57.6 (range 24 to 83) and 60% were males. The second group consisted of adults infected with SARS-CoV-2 who were treated for COVID-19 as outpatients (N=51). This group had a mean age of 36.8 years (range 23-77) and was 25.5% male (Table 1). Differences between these two SARS-CoV-2-infected groups, in terms of median age (p = 5.2 x 10^-8^) and sex ratio (p = 3.7 x 10^-3^) (Fig. 1), were consistent with the established greater risk of severe COVID-19 in older individuals and in males (Alkhouli et al., 2020; Bunders and Altfeld, 2020; Gupta et al., 2021; Laxminarayan et al., 2020; Mauvais-Jarvis, 2020; O’Driscoll et al., 2020; Peckham et al., 2020; Richardson et al., 2020; Scully et al., 2020). Available information concerning ethnicity and race of the blood donors was insufficient for statistical comparisons among the groups (Supplementary Table S1). Finally, 86 adults who donated blood prior to the SARS-CoV-2 outbreak, or who were screened at a blood donation center, were included as controls for SARS-CoV-2 infection. The age of this group spanned the range of the two groups of SARS-CoV-2-infected people (mean age 50.9; range 23 to 79), and the percentage of males (55.8%) was similar to that of the group hospitalized for COVID-19 (Table 1 and Fig. 1).

**Fig. 1.**
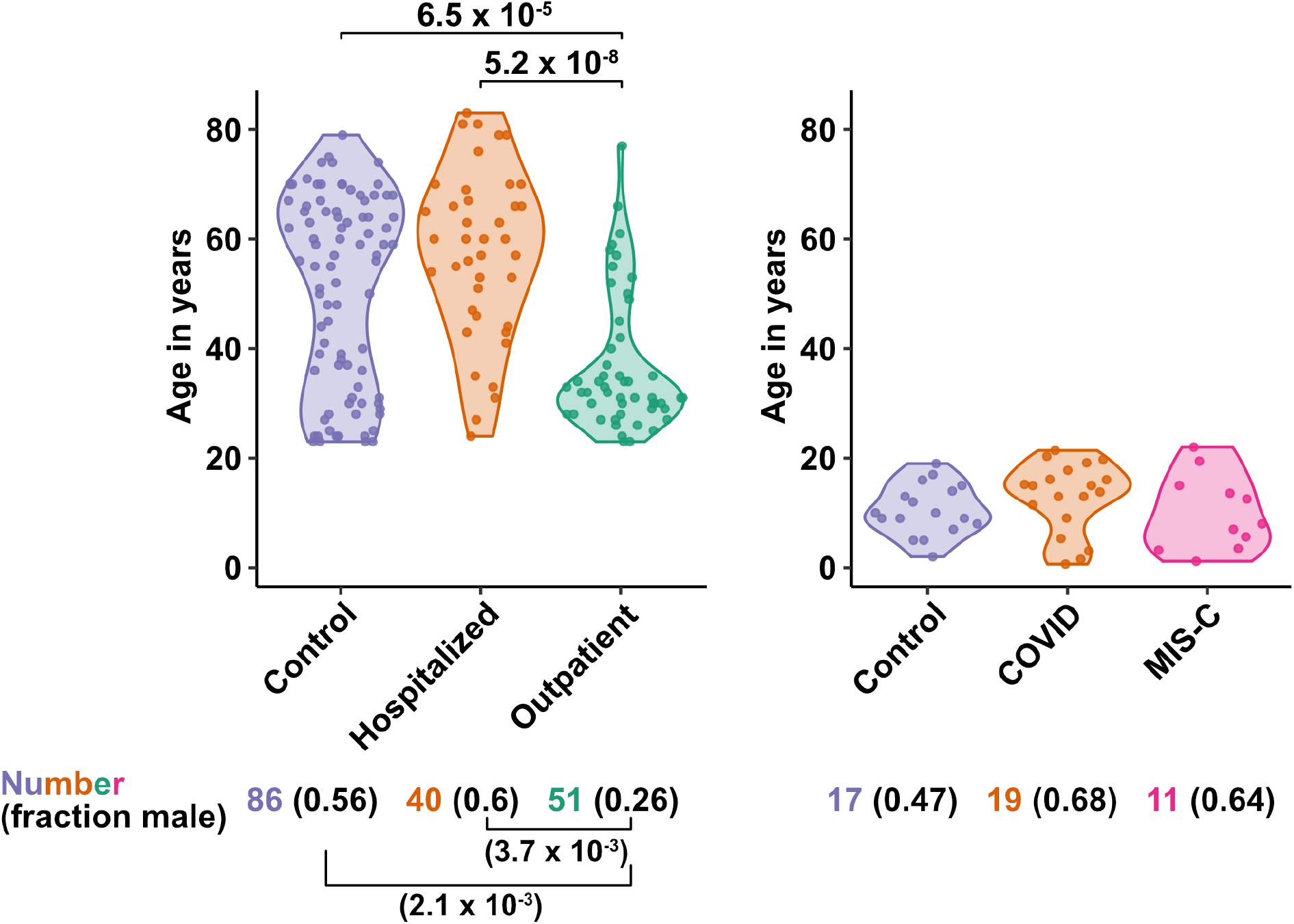
**Age and sex of control and SARS-CoV-2-infected blood donors** Age of the subjects is shown, along with the number of subjects and fraction male in each group, for adult (left) and pediatric (right) cohorts, as indicated. P-values are from pairwise, two-sided, Wilcoxon rank-sum test for ages and Fisher’s exact test for fraction male, with Bonferroni correction for multiple comparisons. Adjusted P-values < 0.05 are shown.

**Table 1:** Demographic and Clinical Characteristics of Adult Blood Donor Groups.

| **Characteristic** | **Healthy Control** | **Hospitalized** | **Outpatient** |
| --- | --- | --- | --- |
|  | **N=86** | **N=40** | **N=51** |
| Mean age (range) - years | 50.9 (23-79) | 57.6 (24-83) | 36.8 (23-77) |
| Sex – number (%) |  |  |  |
| Male | 48 (55.8) | 24 (60) | 13 (25.5) |
| Female | 38 (44.2) | 16 (40) | 38 (74.5) |
| Mean symptom duration at sample collection (range) – days |  | 21.8 (5-66) | 26.9 (1-61) |
| ICU admission – number (%) |  | 33 (82.5) |  |
| Intubation with mechanical ventilation – number (%) |  | 32 (80) |  |
| Deaths – number (%) |  | 7 (17.5) |  |
| Mean time hospitalized (range) – days |  | 34.2 (4-87) |  |
| Max lab value – mean (range) |  |  |  |
| CRP – mg/L |  | 228.6 (6.5-539.5) |  |
| ESR – mm/h |  | 89.0 (15-146) |  |
| D-dimer – (ng/mL) |  | 5700 (351-11923) |  |

### Characteristics of pediatric blood donors with COVID-19, MIS-C, or SARS-CoV-2-uninfected controls

Children are less likely than adults to have severe disease when infected with SARS-CoV-2 despite having viral loads as high as adults (Charles Bailey et al., 2020; Heald-Sargent et al., 2020; Li et al., 2020; LoTempio et al., 2021; Lu et al., 2020; Poline et al., 2020; Yonker et al., 2020). Rarely, after SARS-CoV-2 clearance from the upper airways, children can develop severe Multisystem Inflammatory Syndrome in Children (MIS-C), a life-threatening condition distinct from COVID-19 that presents with high fevers and multiorgan injury, often including coronary aneurysms, ventricular failure, or myocarditis (Cheung et al., 2020; Feldstein et al., 2021, 2020; Licciardi et al., 2020; Riphagen et al., 2020; Verdoni et al., 2020; Whittaker et al., 2020).

The first cohort of pediatric blood donors in this study consisted of patients with COVID-19 who were treated in hospital (N=11) or as outpatients (N=8). The second cohort of pediatric blood donors was patients hospitalized for MIS-C (N=11). Seventeen SARS-CoV-2-uninfected pediatric blood donors constituted a control group. No significant differences in age or percentage of males were detected among the pediatric COVID-19, MIS-C, or pediatric control groups (Supplementary Table S2 and Fig. 1).

### Blood ILC abundance decreases exponentially across the lifespan and is sexually dimorphic

Lymphoid cell abundance in peripheral blood changes with age and is sexually dimorphic (Márquez et al., 2020; Patin et al., 2018). Previous studies reporting the effect of COVID 19 on the abundance of blood lymphoid cell subsets have not fully accounted for the association of age and sex with COVID-19 severity. To isolate the effect of COVID-19 on cell abundance from effects of age and sex, PBMCs were collected from 103 SARS-CoV-2-negative blood donors distributed from 2 to 79 years of age, with a nearly equal ratio of males to females. Abundance of lymphoid cell types was plotted by 20-year age groups (Fig. 2A), as well as by sex (Fig. 2B). Lymphoid cell types assessed here included CD4^+^ T cells, CD8^+^ T cells, ILCs, and FcγRIII (CD16)-positive NK cells. Like CD8^+^ T cells, NK cells kill virus-infected cells using perforin and granzyme (Artis and Spits, 2015; Cherrier et al., 2018). Additionally, by binding virus-specific immunoglobulins that target virus-infected cells for antibody-dependent cellular cytotoxicity, CD16^+^ NK cells link innate and acquired immunity (Anegon et al., 1988).

**Fig. 2.**
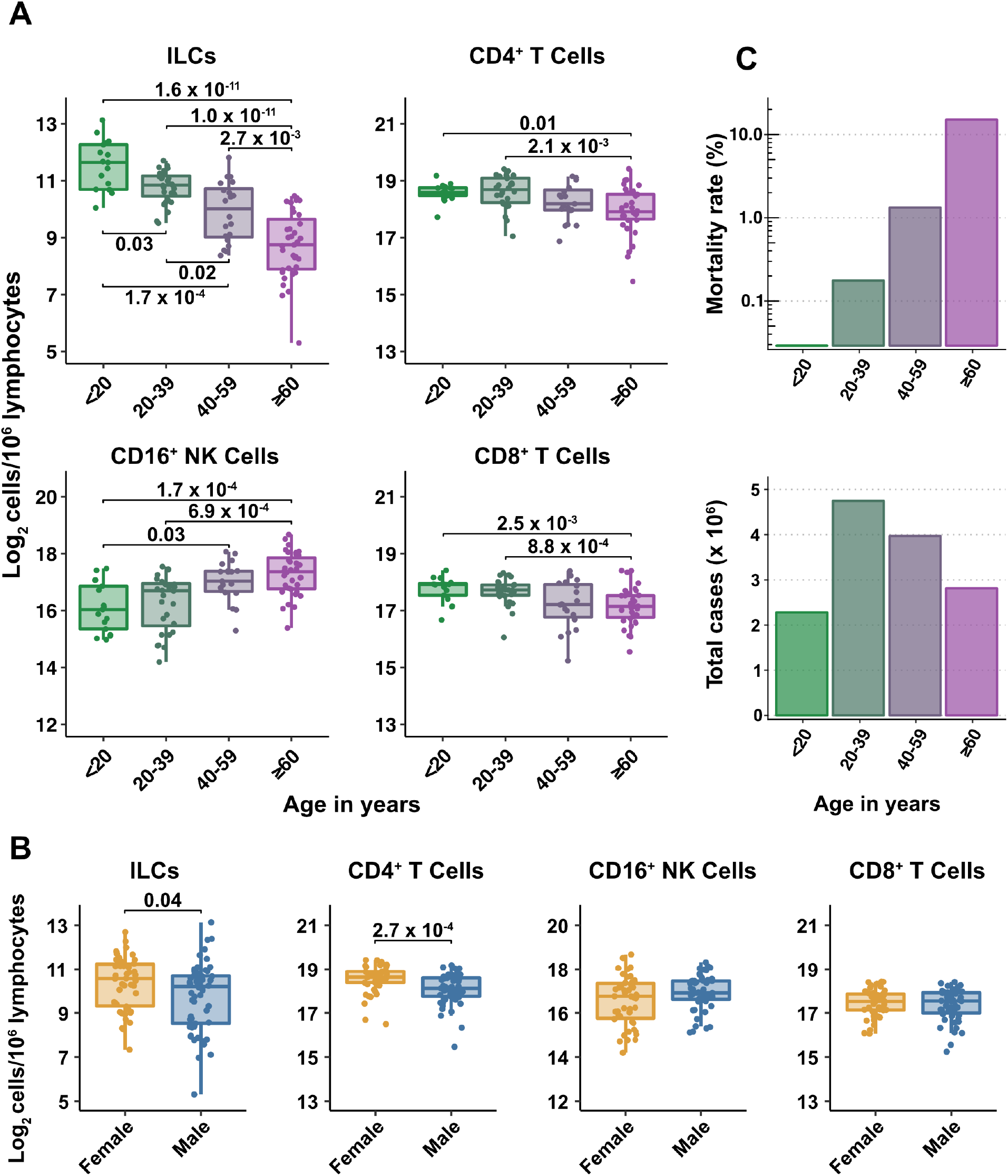
**Blood ILC abundance decreases exponentially across the lifespan mirroring the mortality rate from SARS-CoV-2 infection** (**A-B**) Log2 abundance per million lymphocytes of the indicated lymphoid cell populations in combined pediatric and adult control data plotted by 20-year bin or by sex, as indicated. Each dot represents an individual blood donor. Boxplots represent the distribution of the data with the center line drawn through the median with the upper and lower bounds of the box at the 75th and 25th percentiles respectively. The upper and lower whiskers extend to the largest or smallest values within 1.5 x the interquartile range (IQR). P-values are from two-sided, Wilcoxon rank-sum tests with Bonferroni correction for multiple comparisons. Adjusted P-values < 0.05 are shown. (**C**) Case numbers and mortality rate within the indicated age ranges for cases reported in the United States between Jan 1, 2020, and June 6, 2021.

All cell types examined here were affected by age, but ILCs were the only subset with significant differences among all age groups, falling approximately 2-fold in median abundance every 20 years, with a greater than 7-fold decrease from the youngest to oldest age groups (p = 1.64 x 10^-11^) (Fig. 2A). This magnitude of decrease was unique to ILCs and corresponded inversely with the exponential increase in COVID-19 mortality with age (O’Driscoll et al., 2020) (Fig. 2C). In addition, both ILCs and CD4^+^ T cells were less abundant in males (Fig. 2B). These findings highlight the importance of accounting for effects of age and sex when assessing group differences in lymphoid cell abundance, particularly in the context of a disease such as COVID-19 that disproportionately affects older males (O’Driscoll et al., 2020).

### Adults hospitalized with COVID-19 have fewer total lymphocytes even after accounting for effects of age and sex

Severe COVID-19 is associated with lymphopenia (Chen et al., 2020; Huang et al., 2020; Huang and Pranata, 2020; Zhang et al., 2020; Zhao et al., 2020) but it remains unclear if this effect is due to reduction in particular lymphoid cell subpopulations, or whether this effect is explained by the more advanced age and higher proportion of males among people with severe COVID-19. As a first step to assess the specificity of lymphocyte depletion, the effect of COVID-19 on total lymphocyte abundance was addressed with multiple linear regression. After accounting for effects of age and sex, individuals hospitalized with severe COVID-19 had 1.33-fold (95%CI: 1.49–1.19; p = 1.22 x 10^-6^) fewer total lymphocytes among PBMCs than did controls (Supplementary Table S3). Lymphocyte abundance in people infected with SARS-CoV-2 who were treated as outpatients was not different from controls (Supplementary Table S3). In addition, total lymphocytes decreased with age and were less abundant in males (Supplementary Table S3). Subsequent analyses of lymphoid cell subsets took into account the depletion in total lymphocytes associated with COVID-19 by assessing lymphoid subsets as a fraction of total lymphocytes.

### After accounting for age and sex, only innate lymphoid cells are depleted in severe COVID-19

To determine whether there were independent associations between lymphoid cell subsets and COVID-19, multiple linear regression was performed on the abundance of lymphoid cell subsets, with age, sex, and group (control, hospitalized, and outpatient) as independent variables. Across all three groups of adult blood donors, CD4^+^ T cells, CD8^+^ T cells, and ILCs decreased with age, while CD16^+^ NK cells increased with age, and both CD4^+^ T cells and ILCs were less abundant in males (Table 2 and Fig. 3A).

**Fig. 3.**
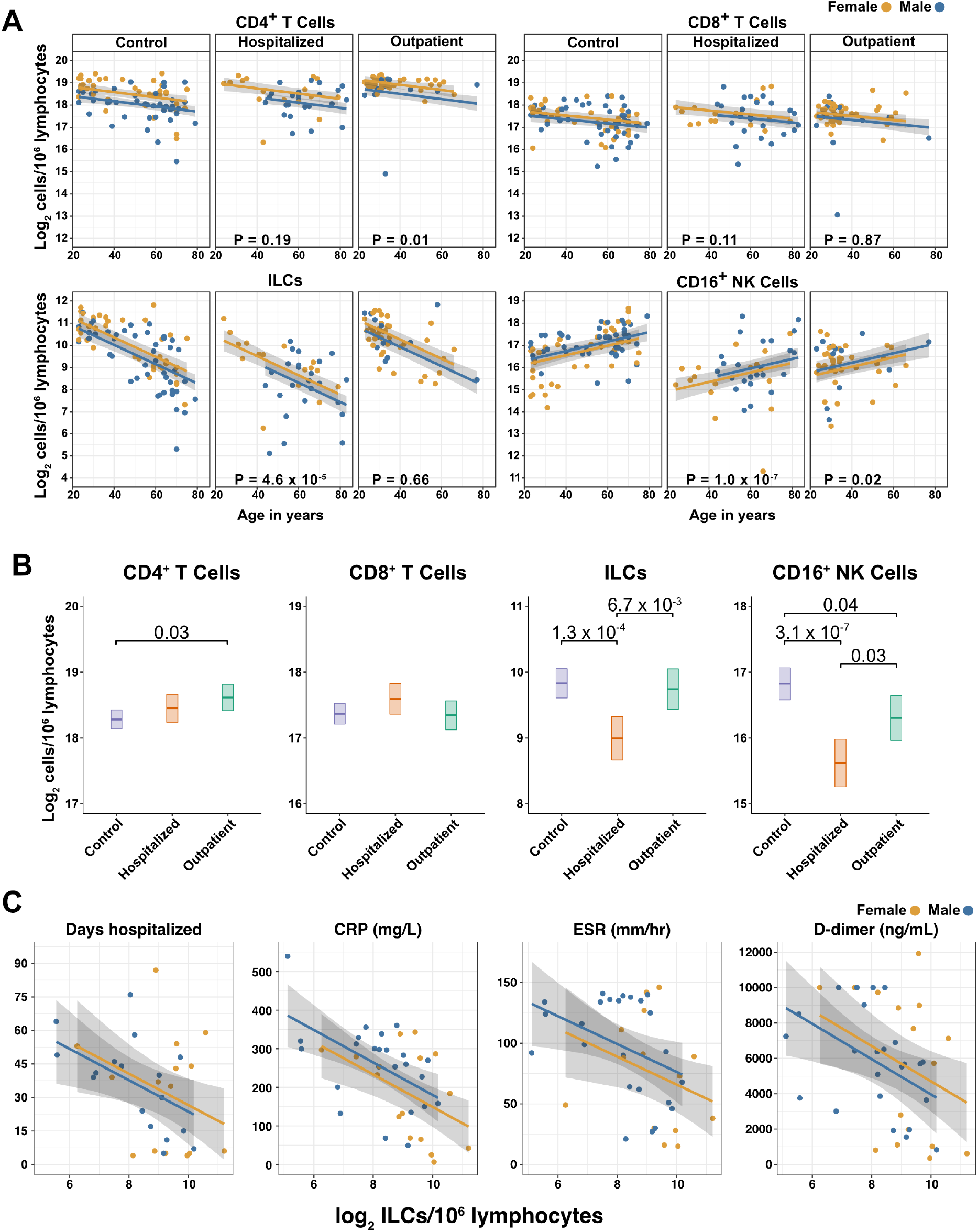
**Innate lymphoid cells are depleted in adults hospitalized with COVID-19 and ILC abundance correlates inversely with disease severity.** (A) Effect of age (X-axis) on log2 abundance per million total lymphocytes of the indicated lymphoid cell populations (Y-axis), as determined by the regression analysis in Table 2. Each dot represents an individual blood donor, with yellow for female and blue for male. Shading represents the 95%CI. P-values are from the regression analysis for comparisons to the control group. (B) Log2 abundance per million lymphocytes of the indicated lymphoid cell populations, shown as estimated marginal means with 95%CI, generated from the multiple linear regressions in Table 2, and averaged across age and sex. P-values represent pairwise comparisons on the estimated marginal means, adjusted for multiple comparisons with the Tukey method. Adjusted P-values < 0.05 are shown. (C) Association of the indicated clinical parameters with log2 abundance of ILCs per million lymphoid cells. Regression lines are from simplified multiple regression models to permit visualization on a two-dimensional plane. Shading represents the 95%CI. Results of the full models accounting for effects of both age and sex, are reported in Table 4 and the text.

**Table 2:** Change in Cell Abundance Due to Age, Sex, and COVID-19 Severity.

Fold difference (log2) [ ± 95%CI]
|  | CD4<sup>+</sup> T<sup>a</sup> | ILC<sup>a</sup> | CD8<sup>+</sup> T<sup>a</sup> | CD16<sup>+</sup> NK<sup>a</sup> |
| --- | --- | --- | --- | --- |
| Age | -0.012***<br>[-0.018, -0.005] | -0.043***<br>[-0.053, -0.033] | -0.009*<br>[-0.016, -0.002] | 0.021***<br>[0.010, 0.032] |
| Male | -0.409***<br>[-0.618, -0.201] | -0.334*<br>[-0.659, -0.010] | -0.177<br>[-0.406, 0.051] | 0.184<br>[-0.169, 0.538] |
| Hospitalized | 0.168<br>[-0.084, 0.421] | -0.835***<br>[-1.228, -0.441] | 0.227<br>[-0.050, 0.503] | -1.205***<br>[-1.633, -0.778] |
| Outpatient | 0.332*<br>[0.082, 0.581] | -0.088<br>[-0.478, 0.302] | -0.023<br>[-0.298, 0.253] | -0.522*<br>[-0.948, -0.095] |
| R<sup>2</sup> | 0.275 | 0.478 | 0.070 | 0.232 |

\* p < 0.05, \*\* p < 0.01, \*\*\* p < 0.001
<sup>a</sup> per 10<sup>6</sup> lymphocytes

When effects of age and sex were held constant, adults hospitalized with COVID-19 had 1.78-fold fewer ILCs (95%CI: 2.34–1.36; p = 4.55 x 10^-5^) and 2.31-fold fewer CD16^+^ natural killer (NK) cells (95%CI: 3.1–1.71; p = 1.04 x 10^-7^), as compared to controls (Table 2 and Fig. 3B). Similar effects were also seen with ILC precursors (ILCP) (Supplementary Fig. S1). Neither CD4^+^ T cells nor CD8^+^ T cells were depleted further than expected for age and sex (Table 2 and Fig. 3B). As compared with controls, SARS-CoV-2-infected adults with less severe COVID-19 who were treated as outpatients had no reduction in ILCs, but 1.44-fold fewer CD16^+^ NK cells (95%CI: 1.93–1.07; p = 0.018), and 1.26-fold higher CD4^+^ T cells (95%CI: 1.06–1.5; p = 9.59 x 10^-3^) (Table 2 and Fig. 3B). As these analyses were performed on lymphoid cell abundance normalized to total lymphocyte number, it is possible that T cells were not lower in patients hospitalized with COVID-19 because the amount of depletion was not in excess of the change in total lymphocytes. However, the cell-type specific results remained unchanged even when the analyses were repeated using the less stringent threshold of normalizing to total PBMC number (Supplementary Table S4).

When data from an independent, previously published cohort (Kuri-Cervantes et al., 2020) were analyzed to account for total lymphocyte abundance, age, and sex, people hospitalized with acute respiratory distress syndrome due to COVID-19, had 1.7-fold fewer ILCs (95%CI: 2.38–1.22; p = 0.002) than controls (Supplementary Fig. S2). Also consistent with the main adult cohort studied here, ILC abundance was not significantly reduced in the group of patients with less severe disease (Supplementary Fig. S2).

### Odds of hospitalization in adults infected with SARS-CoV-2 increases with decreasing number of ILCs

Multiple logistic regression was used next to determine whether differences in abundance of any lymphoid cell subset was associated with odds of hospitalization in people infected with SARS-CoV-2. The adjusted odds ratio was calculated using lymphoid cell subset abundance, age, sex, and duration of symptoms at the time of blood draw, each as independent variables. Abundance of ILCs, but not of CD16^+^ NK cells, CD4^+^ T cells, or CD8^+^ T cells was associated with odds of hospitalization: the odds ratio for hospitalization, adjusted for age, sex, and symptom duration, was 0.413 (95%CI: 0.197–0.724; p = 0.00691), an increase of 58.7% for each 2-fold decrease in ILC abundance (Table 3).

**Table 3:**
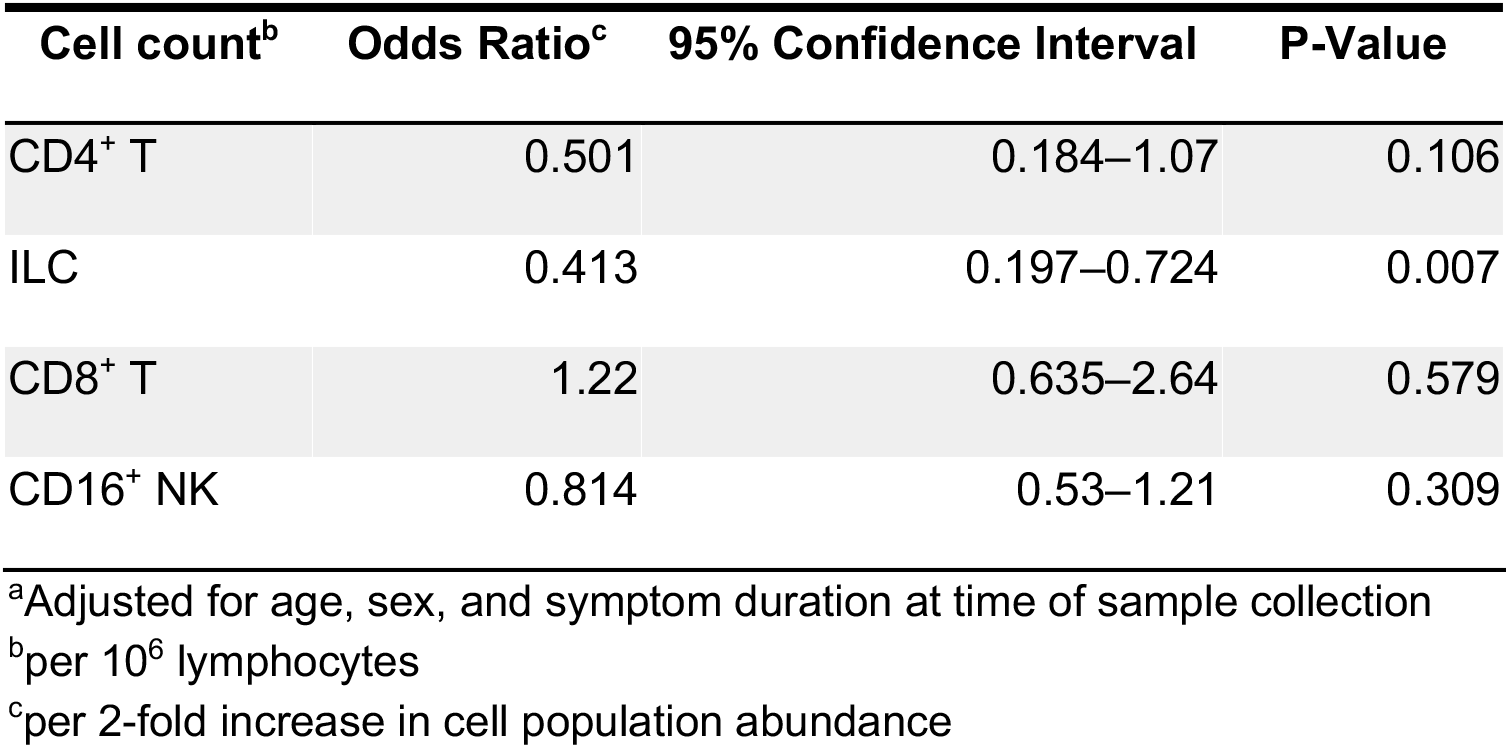
Odds of Hospitalization^a^.

| Cell count<sup>b</sup> | Odds Ratio<sup>c</sup> | 95% Confidence Interval | P-Value |
| --- | --- | --- | --- |
| CD4<sup>+</sup> T | 0.501 | 0.184–1.07 | 0.106 |
| ILC | 0.413 | 0.197–0.724 | 0.007 |
| CD8<sup>+</sup> T | 1.22 | 0.635–2.64 | 0.579 |
| CD16<sup>+</sup> NK | 0.814 | 0.53–1.21 | 0.309 |

<sup>a</sup>Adjusted for age, sex, and symptom duration at time of sample collection
<sup>b</sup>per 10<sup>6</sup> lymphocytes
<sup>c</sup>per 2-fold increase in cell population abundance

**Table 4:**
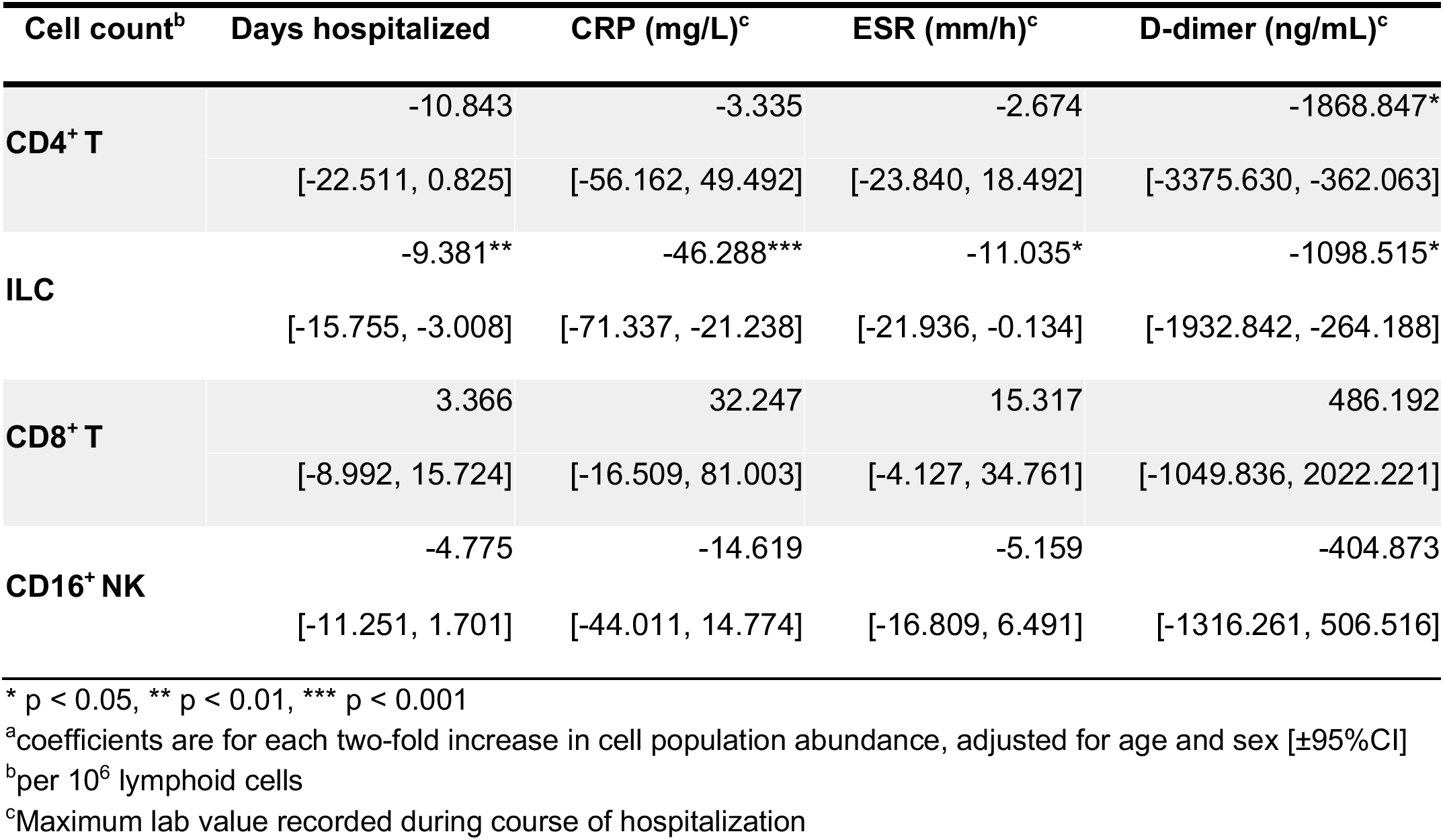
Association of cell type abundance with time in hospital and laboratory values^a^.

### Duration of hospital stay in adults with COVID-19 increases with decreasing ILC abundance

The relationship between lymphoid cell abundance and duration of hospitalization was assessed to determine whether the association between ILC abundance and COVID-19 severity extended to clinical outcomes within the hospitalized adults. This relationship was assessed with multiple linear regression, including age, sex, and cell abundance as independent variables. Holding age and sex constant, abundance of ILCs, but not of CD16^+^ NK cells, CD4^+^ T cells, or CD8^+^ T cells, was associated with length of time in the hospital: each two-fold decrease in ILC abundance was associated with a 9.38 day increase in duration of hospital stay (95% CI: 15.76–3.01; p = 0.0054) (Fig. 3C and Table 4).

### ILC abundance correlates inversely with markers of inflammation in adults hospitalized with COVID-19

To further characterize the extent to which lymphoid cell abundance predicted COVID-19 severity, multiple regression with age, sex, and cell abundance, as independent variables, was performed on peak blood levels of inflammation markers indicative of COVID-19 severity: C-reactive protein (CRP) and erythrocyte sedimentation rate (ESR), and the fibrin degradation product D-dimer (Gallo Marin et al., 2020; Gupta et al., 2021; Luo et al., 2020; Zhang et al., 2020; Zhou et al., 2020). Holding age and sex constant, each two-fold decrease in ILC, but not in CD16^+^ NK cell, CD4^+^ T cell, or CD8^+^ T cell abundance, was associated with a 46.29 mg/L increase in blood CRP (95% CI: 71.34–21.24; p = 6.25 x 10^-4^) and 11.04 mm/h increase in ESR (95% CI: 21.94–0.13; p = 0.047) (Fig. 3C and Table 4). Abundance of both ILCs and CD4^+^ T cells was associated with blood levels of D-dimer, with each two-fold decrease in cell abundance associated with an increase in D-dimer by 1098.52 ng/mL (95% CI: 1932.84–264.19; p = 0.011) and 1868.85 ng/mL (95% CI: 3375.63–362.06; p = 0.016), respectively (Table 4).

### ILCs are depleted in children and young adults with COVID-19 or MIS-C

Given the decline in ILC abundance with age (Fig.s 2A and 3A, and Table 2), and the inverse relationship between ILC abundance and disease severity in adults (Fig. 3C, Table 3, and Table 4), it was hypothesized that children as a group have less severe COVID-19 because ILC abundance is higher at younger ages, and that pediatric cases with symptomatic SARS-CoV-2 infection, or with MIS-C, are accompanied by significantly lower numbers of ILCs. To test these hypotheses, the abundance of lymphoid cell subsets in pediatric COVID-19 or MIS-C was compared with that from pediatric controls, using multiple linear regression with age, sex, and group as independent variables. Consistent with the findings in adults, blood ILCs in the pediatric cohort decreased with age (Table 5 and Fig. 4A), demonstrating that the decrease in ILC abundance across the lifespan is already evident within the first two decades of life. In contrast, significant change over this age range was not detected in the abundance of CD4^+^ T cells, CD8^+^ T cells, or CD16^+^ NK cells (Table 5 and Fig 4A).

**Fig. 4.**
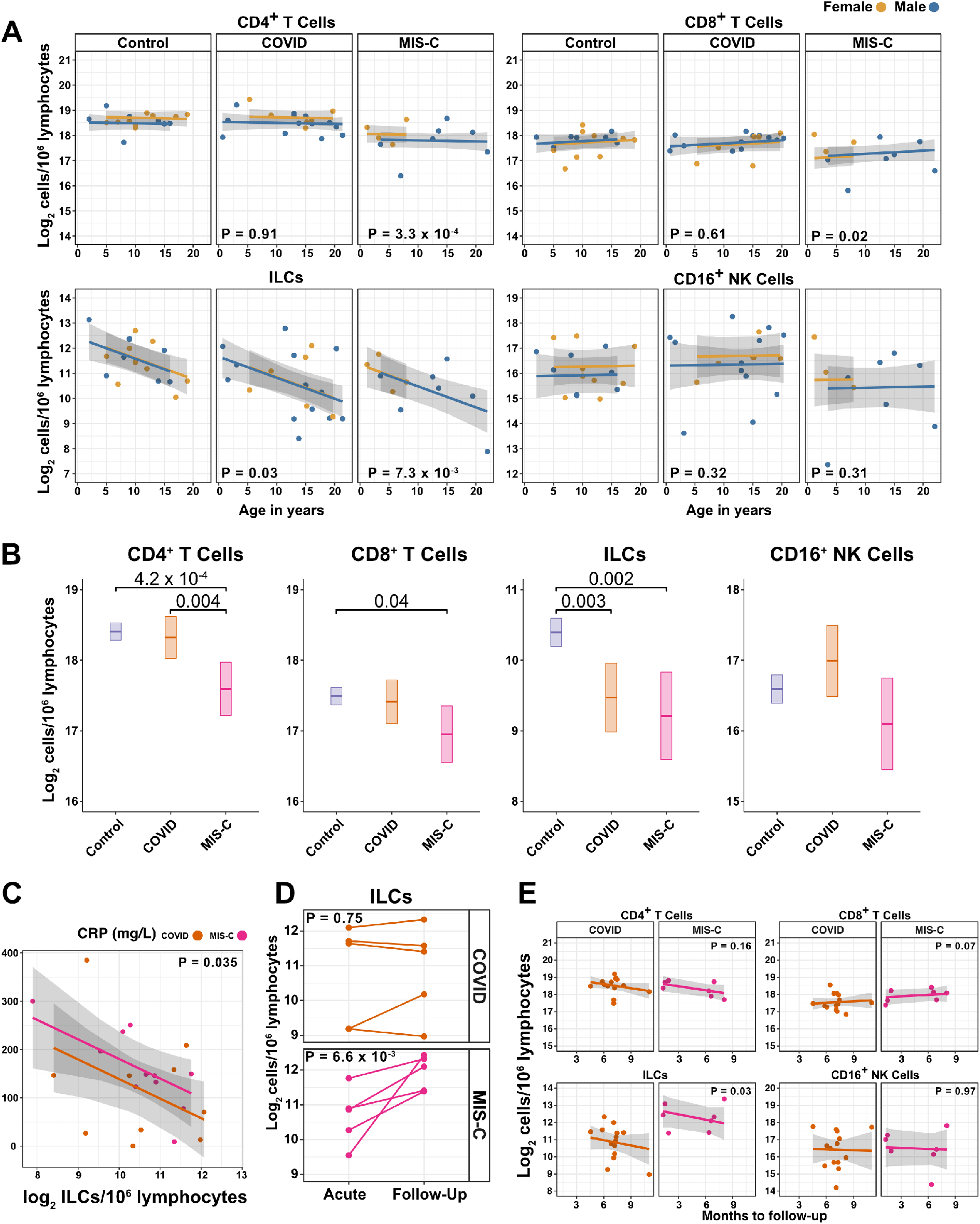
**ILCs are depleted in children with COVID-19 or MIS-C** (A) Effect of age (X-axis) on log2 abundance per million lymphocytes of the indicated lymphoid cell populations (Y-axis), as determined by the regression analysis in Table 5. Each dot represents an individual blood donor, with yellow for female and blue for male. Shading represents the 95%CI. P-values are from the regression analysis for comparisons to the control group. (B) Log2 abundance per million lymphocytes of the indicated lymphoid cell populations, shown as estimated marginal means with 95%CI, generated from the multiple linear regressions in Table S6 that included the combined pediatric and adult control data, and averaged across age and sex. P-values represent pairwise comparisons on the estimated marginal means, adjusted for multiple comparisons with the Tukey method. Adjusted P-values < 0.05 are shown. (C) Association of CRP with log2 abundance of ILCs per million lymphocytes. Shading represents the 95%CI. Each dot represents a single blood donor, orange for COVID-19, magenta for MIS-C. P-value is for the effect of ILC abundance on CRP as determined by linear regression. (D) Log2 ILC abundance per million lymphocytes in longitudinal pairs of samples collected during acute presentation and during follow-up, from individual children with COVID-19 or MIS-C. Each pair of dots connected by a line represents an individual blood donor. P-values are for change in ILC abundance at follow-up, as determined with a linear mixed model, adjusting for age, sex, and group, and with patient as a random effect. (E) Effect of time to follow-up (X-axis) on log2 abundance per million lymphocytes of the indicated lymphoid cell populations (Y-axis). P-values are for the difference between the COVID-19 and MIS-C follow-up groups, independent of time to follow-up as determined by linear regression. Shading represents the 95%CI.

**Table 5:**
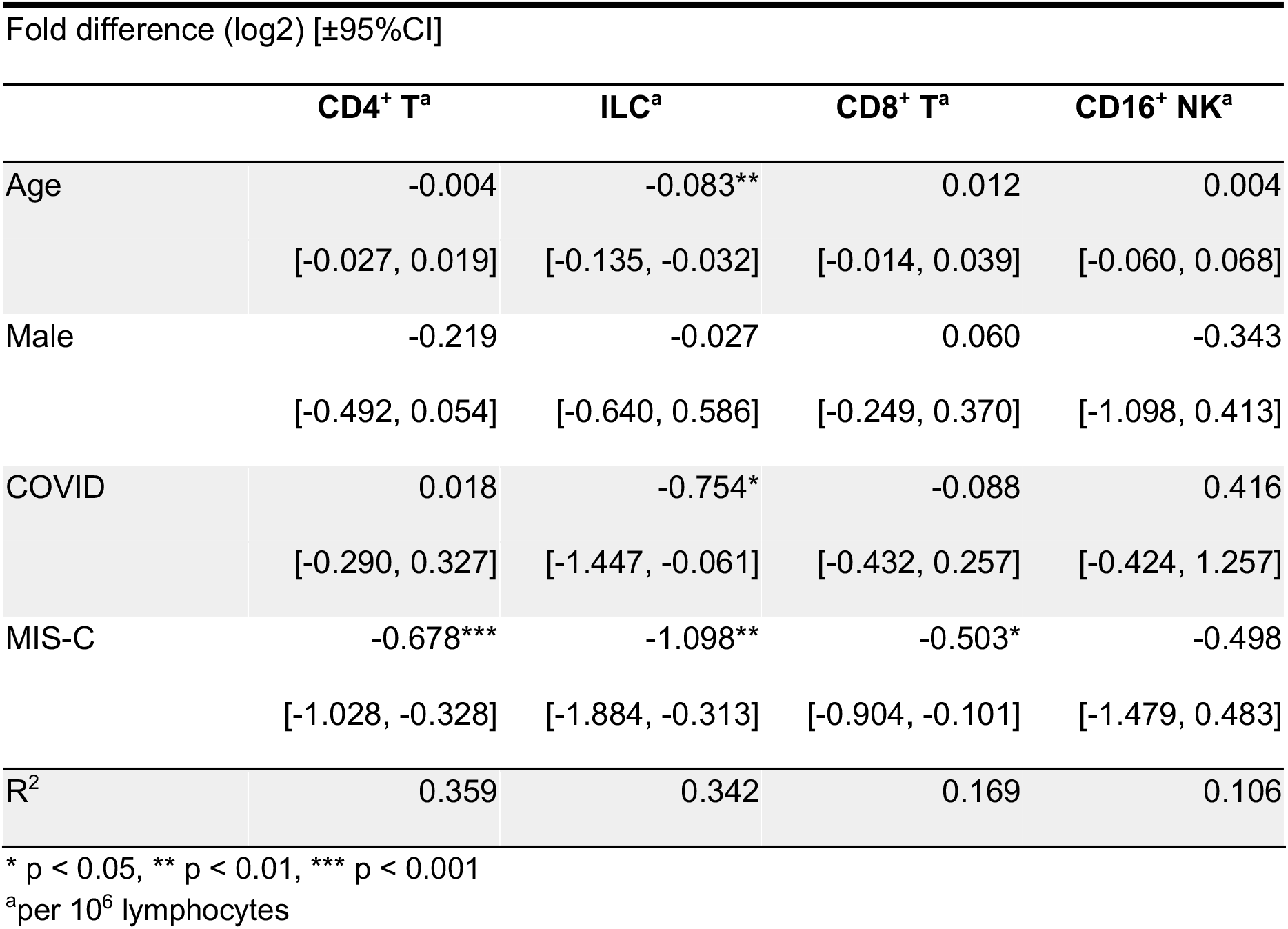
Change in Pediatric Cohort Cell Abundance Due to Age, Sex, and Group.

Among pediatric patients with COVID-19, no difference in abundance of the lymphoid cell subsets was associated with hospitalization (Supplementary Table S5), so all pediatric patients treated for COVID-19 were analyzed as a single group. After accounting for effects of age and sex, pediatric patients with COVID-19 had 1.69-fold fewer ILCs (95%CI: 2.73–1.04; p = 0.034) than controls (Fig 4A and Table 5). Neither CD4^+^ T cells, CD8^+^ T cells, nor CD16^+^ NK cells were depleted in pediatric COVID-19 patients (Fig 4A and Table 5).

As with pediatric COVID-19, ILCs were also lower in MIS-C, with 2.14-fold fewer ILCs (95%CI: 3.69–1.24; p = 0.007) than controls (Fig. 4A and Table 5). However, unlike pediatric COVID-19, individuals with MIS-C had reduced numbers of T cells as compared with pediatric controls, with 1.6-fold fewer CD4^+^ T cells (95%CI: 2.04–1.26; p = 3.28x10^-4^) and 1.42-fold fewer CD8^+^ T cells (95%CI: 1.87–1.07; p = 0.016) (Fig. 4A and Table 5). Depletion of T cells, then, distinguished MIS-C from both pediatric and adult COVID-19. Additionally, consistent with the finding in adults hospitalized with COVID-19 (Fig. 3C and Table 4), after accounting for effect of group, each two-fold decrease in ILC abundance in pediatric patients hospitalized with COVID-19 or MIS-C was associated with a 40.5 mg/L increase in blood CRP (95% CI: 77.87–3.13; p = 0.035) (Fig. 4C), and no such association was detected with CD4^+^ T cells, CD8^+^ T cells, or CD16+ NK cells.

The above analysis of lymphoid cell subsets in pediatric COVID-19 and MIS-C was performed in comparison to pediatric controls alone. Results were essentially unchanged when multiple linear regression was repeated with combined pediatric and adult control groups (Fig. 4B, Supplementary Fig. S3, and Supplementary Table S6).

### Pediatric MIS-C is distinguished from COVID-19 by recovery of ILCs during follow-up

The availability of follow-up samples in this pediatric cohort provided the opportunity to assess the abundance of lymphoid subsets after recovery from illness. To this end, a linear mixed model was fit to determine the change in ILC abundance from acute illness to follow-up in 10 individuals (5 COVID-19 and 5 MIS-C) for whom both acute and follow-up samples were available. After accounting for effects of age, sex, and group, individuals recovering from MIS-C had a 2.39-fold increase in ILC abundance (95%CI: 1.49–3.81; p = 6.6x10^-3^) but there was no significant change in ILC abundance for individuals recovering from COVID-19 (Fig. 4D). Both CD4^+^ and CD8^+^ T cells, which were depleted in MIS-C but not in COVID-19, also increased during recovery from MIS-C and remained unchanged during recovery from COVID-19 (Supplementary Fig. S4).

The relationship between time to follow-up and lymphoid cell abundance was then examined for all available follow-up samples whether or not a paired sample from the acute illness was available (COVID-19, N=14; MIS-C, N=7). This analysis found no relationship between time to follow-up and abundance of any lymphoid subset, and that individuals recovering from MIS-C had 2.28-fold more ILCs (95%CI: 1.11–4.69; p = 0.0265) than individuals recovering from COVID-19 (Fig. 4E). There was no difference between the follow-up groups in CD4^+^ T cells, CD8^+^ T cells, or CD16^+^ NK cells (Fig. 4E). Interestingly, prior to being hospitalized with MIS-C, only one of these patients had COVID-19 symptoms and, despite low ILC abundance in the COVID-19 follow-up cohort, only 28.6% of this group had been ill enough to require hospitalization (Supplementary Table S2).

Differences between COVID-19 and MIS-C in regards to T cell depletion and ILC recovery during follow-up indicate that the underlying processes causing lower ILC abundance in these two SARS-CoV-2-associated diseases are different.

### Blood ILCs resemble homeostatic ILCs isolated from lung

In response to the observations described above regarding abundance of blood ILCs and severity of COVID-19 attempts were made to extend these studies to lung ILCs. It was not possible to obtain lung samples from people with COVID-19. However, ILCs circulate from tissues to the bloodstream in lymphatic drainage via the thoracic duct (Buggert et al., 2020) suggesting that measurement of blood ILCs could provide a surrogate for assessment of tissue-resident ILCs, including those from the lung. Furthermore, reduction in blood ILCs in people living with HIV-1 infection is paralleled by decreased ILC numbers within the lamina propria of the colon (Wang et al., 2020), and so the lower abundance of blood ILCs associated with severe COVID-19 might be paralleled by decreased abundance of homeostatic ILCs in the lung.

Given the inability to assess lung samples from people with COVID-19, RNA sequencing (RNA-Seq) was performed on blood ILCs from nine healthy controls and these data were compared to previously published RNA-Seq profiles of ILCs sorted from lung, spleen, and intestine (Ardain et al., 2019; Yudanin et al., 2019). Unbiased principal component analysis demonstrated overlap of blood ILCs with lung ILCs and clear separation from ILCs of jejunum or spleen (Fig. 5A). 355 genes were consistently differentially expressed (fold-change > 1.5, padj < 0.01) when either blood or lung ILCs were compared to ILCs from the other tissues (Fig. 5B,C). Gene ontology analysis demonstrated enrichment for terms associated with type 2 immunity (Supplementary Table S7). Genes significantly higher in both blood and lung ILCs included the ILC2-defining genes GATA3 and PTGDR2 (CRTH2), as well as other genes important for ILC development such as TCF7 (Yang et al., 2013) (Fig. 5B,C). As confirmation of the RNA signature, TCF7- and CRTH2-encoded proteins were detected in blood ILCs by flow cytometry (Fig. 6A).

**Fig. 5.**
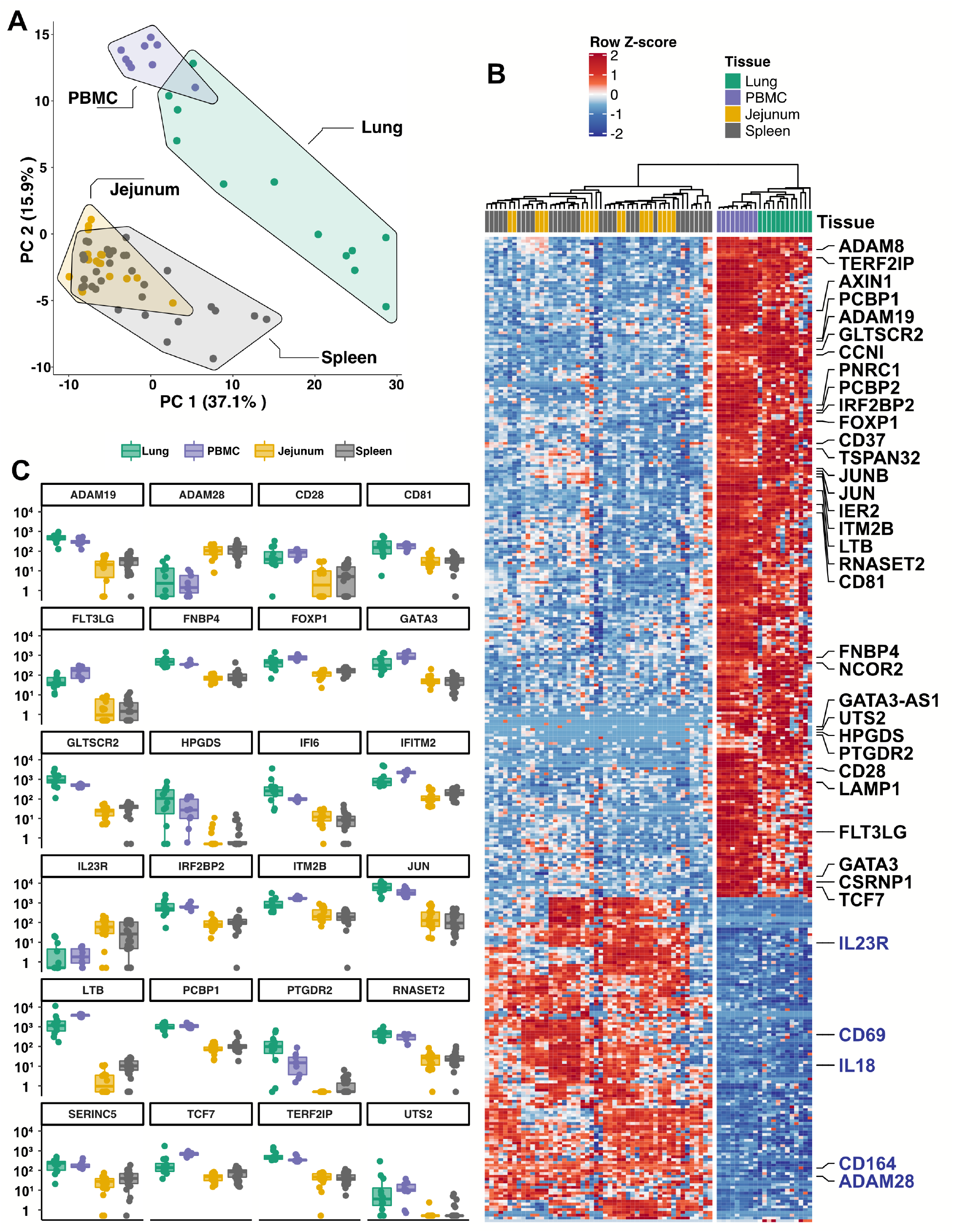
**Blood ILCs are transcriptionally similar to lung ILCs** RNA-seq of ILCs sorted from blood of 9 SARS-CoV-2-uninfected controls in comparison to RNA-seq data of ILCs sorted from jejunum, lung, and spleen. (A) PCA plot of first two principal components calculated from the top 250 most variable genes across all samples. Each dot represents an individual sample with blue for ILCs sorted from blood, green for lung, yellow for jejunum, and grey for spleen. (B) Heatmap of 355 genes differentially expressed (fold-change > 1.5, padj < 0.01 as determined with DESeq2) between either blood or lung ILCs and ILCs from the other tissues. (C) Select genes from (B) plotted as deseq2 normalized counts. Each dot represents an individual sample with blue for ILCs sorted from blood, green for lung, yellow for jejunum, and grey for spleen. Boxplots represent the distribution of the data with the center line drawn through the median with the upper and lower bounds of the box at the 75th and 25th percentiles respectively. The upper and lower whiskers extend to the largest or smallest values within 1.5 x the interquartile range (IQR).

**Fig. 6.**
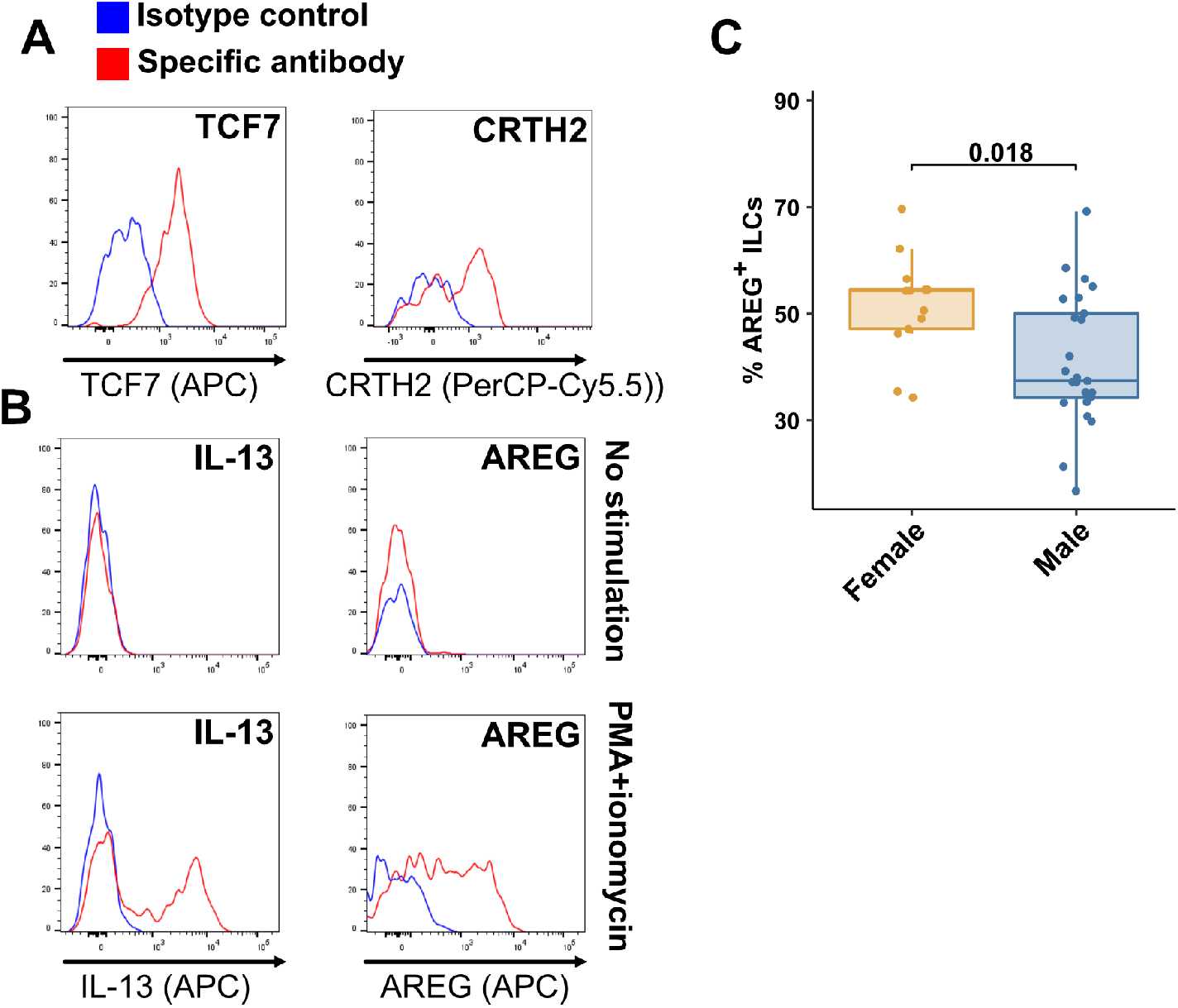
**Peripheral blood ILCs exhibit homeostatic ILC2 functions** (**A-B**) Flow cytometry for the indicated proteins. Cells in (A) were assayed at steady-state and cells in (B) were assayed either at steady-state or after stimulation with PMA and ionomycin, as indicated. Detection of surface proteins was performed on ILCs gated as Lin^-^CD56^-^CD127^+^ and detection of intracellular proteins was performed on ILCs gated as Lin^-^TBX21^-^CD127^+^. (**C**) Percent of AREG^+^ ILCs in blood of control blood donors after stimulation with PMA and ionomycin. Each dot represents an individual blood donor. Boxplots represent the distribution of the data with the center line drawn through the median with the upper and lower bounds of the box at the 75th and 25th percentiles respectively. The upper and lower whiskers extend to the largest or smallest values within 1.5 x the interquartile range (IQR). P-value is from a two-sided, Wilcoxon rank-sum test.

### Blood ILCs are functional ILC2s capable of producing AREG

To assess the function of blood ILCs, PBMCs were stimulated with PMA and ionomycin, and assayed by flow cytometry for production of IL-13 after intracellular cytokine staining and gating on ILCs. IL-13 was detected in the stimulated ILC population (Fig. 6B), demonstrating that the majority of blood ILCs function as ILC2s. Additionally, the blood ILCs produced amphiregulin (Fig. 6B), a protein implicated in the promotion of disease tolerance by ILCs in animal models (Branzk et al., 2018; Diefenbach et al., 2020; Jamieson et al., 2013; McCarville and Ayres, 2018; Monticelli et al., 2015, 2011).

### Females have a higher fraction of amphiregulin-producing blood ILCs than do males

Given the role AREG-producing ILCs play in maintaining disease tolerance in animal models (Branzk et al., 2018; Diefenbach et al., 2020; McCarville and Ayres, 2018; Monticelli et al., 2015, 2011), sex differences in the functional capability of these ILCs could contribute to the greater risk for severe COVID-19 in males (O’Driscoll et al., 2020). To address this hypothesis, ILCs isolated from peripheral blood of controls were stimulated with PMA and ionomycin, and assayed by flow cytometry for AREG production. Consistent with the apparently lower disease tolerance in males, males had a lower median fraction of AREG^+^ ILCs than did females (P=0.018) (Fig. 6C). This difference was also reflected in a significantly lower AREG Mean Fluorescent Intensity (MFI) in males, and neither fraction of AREG^+^ ILCs nor AREG MFI was affected by age (Supplementary Fig. S5).

## DISCUSSION

The outcome of SARS-CoV-2 infection ranges from entirely asymptomatic to lethal COVID-19 (Cevik et al., 2021; He et al., 2021; Jones et al., 2021; Lee et al., 2020; Lennon et al., 2020; Ra et al., 2021; Richardson et al., 2020; Yang et al., 2021). Yet, viral load does not reliably discriminate asymptomatic from symptomatic or hospitalized populations (Cevik et al., 2021; Jones et al., 2021; Lee et al., 2020; Lennon et al., 2020; Ra et al., 2021; Yang et al., 2021). In contrast, demographic factors, including increasing age and male sex, predict worse outcome of SARS-CoV-2 infection (Alkhouli et al., 2020; Bunders and Altfeld, 2020; Gupta et al., 2021; Laxminarayan et al., 2020; Mauvais-Jarvis, 2020; O’Driscoll et al., 2020; Peckham et al., 2020; Richardson et al., 2020; Scully et al., 2020). These demographic risk factors could be due to sexual dimorphism and changes with aging in composition and function of the human immune system (Darboe et al., 2020; Klein and Flanagan, 2016; Márquez et al., 2020; Patin et al., 2018; Solana et al., 2012). Therefore it is necessary to account for effects of age and sex to determine if there are additional, independent, effects of SARS-CoV-2-associated disease.

This study collected and analyzed 245 blood samples from 177 adult and 58 pediatric patients and controls, spanning the ages of 0.7 to 83 years, with approximately equal numbers of males and females. It was therefore possible to characterize the independent effects of age, sex, COVID-19, and MIS-C on blood lymphoid cell populations. After accounting for effects of age and sex, ILCs, but not CD4^+^ or CD8^+^ T cells, were lower in individuals hospitalized with COVID-19 when compared with controls (Table 2 and Fig. 3A,B). Lower numbers of ILCs were also observed in children with COVID-19 (Table 5 and Fig. 4A,B), as well as in an independent cohort of adult patients (Supplementary Fig. S2). Among adults infected with SARS-CoV-2, lower abundance of ILCs, but not of the other lymphoid cell subsets, was associated with increased odds of hospitalization, longer duration of hospitalization, and higher blood level of factors associated with systemic inflammation, including CRP (Tables 3 and 4, and Fig. 3C). This inverse relationship between ILC abundance and CRP was also evident in children with COVID-19 or MIS-C (Fig. 4C).

The identification of reduced ILC numbers as uniquely related to COVID-19 severity is important as these cells mediate disease tolerance in animal models (Artis and Spits, 2015; Branzk et al., 2018; Califano et al., 2018; Diefenbach et al., 2020; McCarville and Ayres, 2018; Monticelli et al., 2015, 2011). The results here therefore indicate that loss of ILCs from blood correlates with loss of ILC-associated homeostatic functions, thereby allowing more severe COVID-19. Although this study examined circulating blood lymphoid cells, and does not provide direct information about processes occurring within tissues, transcriptional and functional characterization of blood ILCs demonstrated that these cells are similar to ILCs isolated from lung tissue (Fig. 5). Human ILCs circulate in lymphatic fluid draining from the tissues to the blood via the thoracic duct (Buggert et al., 2020), raising the possibility that some ILCs in the blood originate from, or traffic to, lung tissue. Further characterization of these blood ILCs showed that they are functional ILC2s capable of producing the protein AREG (Fig. 6A,B). Given the tissue homeostatic role AREG plays in animal models of disease tolerance (Branzk et al., 2018; Diefenbach et al., 2020; Jamieson et al., 2013; McCarville and Ayres, 2018; Monticelli et al., 2015, 2011), the discovery here that males have a smaller fraction than females of blood ILCs capable of producing AREG (Fig. 6C) could explain why males are at greater risk of death from SARS-CoV-2 infection (O’Driscoll et al., 2020). This sexual dimorphism in ILC function would be amplified further by the lower overall abundance of ILCs in males (Fig. 2B and Table 3).

Although the inverse relationship between the number of blood ILCs and severity of COVID-19 suggests that loss of ILC homeostatic function results in breakdown of disease tolerance (Arpaia et al., 2015; Artis and Spits, 2015; Branzk et al., 2018; Diefenbach et al., 2020; McCarville and Ayres, 2018; Monticelli et al., 2015, 2011), this observational study cannot determine whether ILC depletion preceded SARS-CoV-2 infection or whether ILC numbers are depleted as a consequence of SARS-CoV-2 infection. However, several observations support the hypothesis that individuals with lower ILC numbers at the time of SARS-CoV-2 infection are at greater risk of developing severe disease. ILC numbers in uninfected controls decrease exponentially with age; this decrease is much larger than that seen with other lymphoid cell types (Fig. 2A), and much more closely mirrors the exponential increase in COVID-19 mortality with age (O’Driscoll et al., 2020) (Fig. 2C). In addition, the greater risk of COVID-19 mortality in males (O’Driscoll et al., 2020) correlates with lower abundance of blood ILCs (Fig. 2B and Table 3) and smaller fraction of ILCs capable of producing AREG (Fig. 6C). Further supporting this hypothesis is the observation that conditions independently associated with lower ILC abundance, such as HIV-1 infection (Kløverpris et al., 2016; Wang et al., 2020) and obesity (Brestoff et al., 2015; Yudanin et al., 2019), increase the risk for worse outcomes from SARS-CoV-2 infection (Biccard et al., 2021; Kompaniyets, 2021; Tesoriero et al., 2021).

In contrast to individuals with COVID-19, children with MIS-C had lower numbers of T cells as well as ILCs (Table 5 and Fig. 4A,B), and longitudinal follow-up samples for pediatric COVID-19 and MIS-C patients showed persistence of low ILC numbers after COVID-19, but normalization of all depleted cell types after recovery from MIS-C (Fig. 4D,E and Supplementary Fig. S4). These differences imply that the reversible lymphopenia in MIS-C is due to different underlying processes than the more specific and persistent lower ILC abundance seen in individuals with COVID-19. This difference is made more interesting by the fact that none of the children with MIS-C had required hospitalization for COVID-19 and only one experienced any COVID-19 symptoms. The other children with MIS-C were therefore unaware that they had been infected. It is possible that children with pre-existing lower ILC numbers are at risk of developing COVID-19 if infected with SARS-CoV-2, while other factors such as prolonged exposure to SARS-CoV-2 antigens in the gastrointestinal tract (Yonker et al., 2021), or rare inborn errors of immunity (Sancho-Shimizu et al., 2021), promote inflammatory processes in MIS-C that drive nonspecific lymphoid cell depletion, which ultimately normalizes after recovery.

Although ILC depletion and recovery has been reported in rheumatoid arthritis (Rauber et al., 2017), inflammation-driven ILC-depletion is not necessarily reversible, as ILCs appear permanently depleted after HIV-1 infection, possibly by high levels of common γ-chain cytokines during acute infection (Wang et al., 2020). Better understanding of the processes that drive down ILC abundance in populations susceptible to COVID-19 could potentially allow for development of interventions that increase ILC abundance and restore homeostatic disease tolerance mechanisms.

In conclusion, considering the established homeostatic function of ILCs (Artis and Spits, 2015; Branzk et al., 2018; Klose and Artis, 2016; Monticelli et al., 2015, 2011) and presumed non-immunologic, host adaptive responses necessary to survive pathogenic infection (López-Otín and Kroemer, 2021; McCarville and Ayres, 2018; Medzhitov et al., 2012; Schneider and Ayres, 2008), the findings reported here support the hypothesis that loss of disease tolerance mechanisms attributable to ILCs increase the risk of morbidity and mortality with SARS-CoV-2 infection. The findings of this observational study warrant establishment of prospective cohorts to determine whether abundance of ILCs or of other lymphoid cell subsets associated with disease tolerance (Arpaia et al., 2015; Artis and Spits, 2015; Branzk et al., 2018; Diefenbach et al., 2020; McCarville and Ayres, 2018; Monticelli et al., 2015, 2011), predict clinical outcome for infection with SARS-CoV-2 or other lethal pathogens. Understanding the mechanisms that allow an individual to tolerate high-level viral replication without experiencing symptoms, and how these mechanisms can fail and thereby allow for progression to severe disease, will provide the foundation for development of therapeutic interventions that maintain health and improve survival of pathogenic viral infection (Ayres, 2020b).

## Materials and Methods

### Data availability

The data that support the findings of this study are available within the manuscript and in its supplementary information files. Bulk RNA-seq datasets generated here can be found at: NCBI Gene Expression Omnibus (GEO): GSE168212. Bulk RNA-seq data generated by previously-published studies are available from NCBI GEO: GSE131031 and GSE126107. This study did not generate unique code. Any additional information required to reanalyze the data reported in this paper is available from the lead contact upon request.

### Peripheral blood PBMCs

As part of a COVID-19 observational study, peripheral blood samples were collected from 91 adults with SARS-CoV-2 infection at the Massachusetts General Hospital and affiliated outpatient clinics. Request for access to coded patient samples was reviewed by the Massachusetts Consortium for Pathogen Readiness (https://masscpr.hms.harvard.edu/), and approved by the University of Massachusetts Medical School IRB (protocol #H00020836). Pediatric participants with COVID-19 or MIS-C were enrolled in the Massachusetts General Hospital Pediatric COVID-19 Biorepository (MGB IRB # 2020P000955); healthy pediatric controls were enrolled in the Pediatric Biorepository (MGB IRB # 2016P000949). Samples were collected after obtaining consent from the patient if 18 years or older, or from the parent/guardian, plus assent when appropriate. Demographic, laboratory, and clinical outcome data were included with the coded samples. Samples from 86 adult blood donors and 17 pediatric blood donors, either collected prior to the SARS-CoV-2 outbreak, or from healthy individuals screened at a blood bank, were included as controls.

### Human mononuclear cell isolation

Human peripheral blood was diluted in an equal volume of RPMI-1640 (Gibco), overlaid on Lymphoprep (STEMSELL, 07851), and centrifuged at 500 x g at room temperature for 30 minutes. Mononuclear cells were washed 3 times with MACS buffer (0.5% BSA and 2 mM EDTA in PBS) and frozen in FBS containing 10% DMSO.

### Flow cytometry

Peripheral blood mononuclear cells (PBMCs) were first stained with Live and Dead violet viability kit (Invitrogen, L-34963). To detect surface molecules, cells were stained in MACS buffer with antibodies (Supplementary Table S8) for 30 min at 4°C in the dark. To detect IL-13 or AREG, cells were stimulated with PMA and ionomycin (eBioscience, 00-4970-03) for 3 hours with Brefeldin A and Monensin (eBioscience, 00-4980-03) present during the stimulation. To detect transcription factors or cytokines, cells were fixed and permeabilized using Foxp3 staining buffer kit (eBioscience, 00-5523-00), then intracellular molecules were stained in permeabilization buffer with antibodies. Cells were detected on a BD Celesta flow cytometer using previously established gating strategies (Wang et al., 2020). Cell subsets were identified using FlowJo^TM^ software (Becton, Dickson and Company). Representative gating strategies are shown in Fig. S6.

### Bulk RNA-Seq Library preparation of PBMC ILCs

The sequencing libraries were prepared using CEL-Seq2 (Hashimshony et al., 2016). RNA from sorted cells was extracted using TRIzol reagent (ThermoFisher, 15596018). 10 ng RNA was used for first strand cDNA synthesis using barcoded primers (the specific primers for each sample were listed in Supplementary Table S9). The second strand was synthesized by NEBNext Second Strand Synthesis Module (NEB, E6111L). The pooled dsDNA was purified with AMPure XP beads (Beckman Coulter, A63880), and subjected to in vitro transcription (IVT) using HiScribe T7 High Yield RNA Synthesis Kit (NEB, E2040S), then treated with ExoSAP-IT (Affymetrix, 78200). IVT RNA was fragmented using RNA fragmentation reagents (Ambion), and underwent another reverse transcription step using random hexamer RT primer-5’-GCC TTG GCA CCC GAG AAT TCC ANN NNN N-3’ to incorporate the second adapter. The final library was amplified with indexed primers: RP1 and RPI1 (Supplementary Table S9), and the bead purified library was quantified with 4200 TapeStation (Agilent Technologies), and paired-end sequenced on Nextseq 500 V2 (Illumina), Read 1: 15 cycles; index 1: 6 cycles; Read 2: 60 cycles.

### RNA-seq analyses

Pooled reads from PBMC ILCs were separated by CEL-Seq2 barcodes, and demultiplexed reads from RNA-seq of ILCs from lung (Ardain et al., 2019), spleen and intestine (Yudanin et al., 2019), were downloaded from GSE131031 and GSE126107. Within the DolphinNext RNA-seq pipeline (Revision 4) (Yukselen et al., 2020), reads were aligned to the hg19 genome using STAR (version 2.1.6) (Dobin et al., 2013) and counts of reads aligned to RefSeq genes were quantified using RSEM (version 1.3.1) (Li and Dewey, 2011). Normalized transcript abundance in the form of TPMs were used to filter out low abundance transcripts with an average of <3 TPMs across libraries. RSEM-generated expected counts were normalized and differential analysis was performed using DEseq2 (Love et al., 2014) in R, with significant genes defined as a greater than 1.5-fold difference and an adjusted p-value <0.01. GO Enrichment Analysis was performed in R using the enrichGO function in the clusterProfiler R package (Yu et al., 2012). Data were transformed using vsd within DEseq2 both for the heatmap visualization with ComplexHeatmap (Gu et al., 2016) and for principal component analysis (PCA) with prcomp on the top 250 most variable genes. Normalized counts were generated for plotting using the counts command in Deseq2.

### Statistical analysis and data visualization

Data were prepared for analysis with tidyverse packages (Wickham et al., 2019) and visualized using the ggplot2 (Wickham, 2016), ggpubr (Kassambara, 2020), and ComplexHeatmap (Gu et al., 2016) packages, within the R computer software environment (version 4.0.2) (R Core Team, 2020). Group differences were determined with pairwise, two-sided, Wilcoxon rank-sum tests, or Fisher’s exact test, as indicated, with Bonferroni correction for multiple comparisons. Multiple linear regression analyses were performed with dependent and independent variables as indicated in the text, using the lm function in R. Pairwise group comparisons on estimated marginal means generated from multiple linear regression were performed using the emmeans package (Lenth, 2020) in R, with multiple comparison correction using the Tukey adjustment. Multiple logistic regressions were performed using the glm function in R. Longitudinal follow-up analyses on pediatric COVID-19 and MIS-C was performed with linear mixed-effect models using lme4 (Bates et al., 2015) in R with the equation: log2 (lymphoid cell abundance) ∼ Age + Sex + Group + Group:Follow_up + (1|Patient_ID). This model tested the effect of followup on ILC abundance in the pediatric COVID-19 and MIS-C groups while accounting for age, sex, and group. Statistical significance was determined with lmerTest (Kuznetsova et al., 2017) in R, using the Satterthwarte’s degrees of freedom method. p<0.05 was considered significant. United States SARS-CoV-2 infection and mortality data were downloaded from (CDC Case Surveillance Task Force, 2020) and cases with age group and outcome available were plotted by age group as indicated. Mortality rate was calculated by dividing the number of fatal cases by the total number of cases with known outcome in each age group as indicated.

## Data Availability

The data that support the findings of this study are available within the manuscript and in its supplementary information files. Bulk RNA-seq datasets generated here can be found at: NCBI Gene Expression Omnibus (GEO): GSE168212. Bulk RNA-seq data generated by previously-published studies are available from NCBI GEO: GSE131031 and GSE126107 (Ardain et al., 2019; Yudanin et al., 2019). This study did not generate unique code. Any additional information required to reanalyze the data reported in this paper is available from the lead contact upon request.

## ACKNOWLEDGEMENTS

We thank the Massachusetts Consortium for Pathogen Readiness Specimen Collection and Processing Team listed below and members of the Yu and Luban Labs. This work was supported in part by the Massachusetts Consortium for Pathogen Readiness through grants from the Evergrande Fund and NIH grants R37AI147868 and R01AI148784 to J.L. and Ruth L. Kirschstein NRSA Fellowship F30HD100110 to N.J.S. The MGH/MassCPR COVID biorepository was supported by a gift from Ms. Enid Schwartz, by the Mark and Lisa Schwartz Foundation, the Massachusetts Consortium for Pathogen Readiness, and the Ragon Institute of MGH, MIT and Harvard. The Pediatric COVID-19 Biorepository was supported by the National Heart, Lung, and Blood Institute (5K08HL143183 to LMY), and the Department of Pediatrics at Massachusetts General Hospital *for* Children (to LMY).

## MGH COVID-19 Collection & Processing Team participants Collection Team

Kendall Lavin-Parsons^1^, Blair Parry^1^, Brendan Lilley^1^, Carl Lodenstein^1^, Brenna McKaig^1^, Nicole Charland^1^, Hargun Khanna^1^, Justin Margolin^1^

Processing Team: Anna Gonye^2^, Irena Gushterova^2^, Tom Lasalle^2^, Nihaarika Sharma^2^, Brian C. Russo^3^, Maricarmen Rojas-Lopez^3^, Moshe Sade-Feldman^4^, Kasidet Manakongtreecheep^4^, Jessica Tantivit^4^, Molly Fisher Thomas^4^

Massachusetts Consortium on Pathogen Readiness: Betelihem A. Abayneh^5^, Patrick Allen^5^, Diane Antille^5^, Katrina Armstrong^5^, Siobhan Boyce^5^, Joan Braley^5^, Karen Branch^5^, Katherine Broderick^5^, Julia Carney^5^, Andrew Chan^5^, Susan Davidson^5^, Michael Dougan^5^, David Drew^5^, Ashley Elliman^5^, Keith Flaherty^5^, Jeanne Flannery^5^, Pamela Forde^5^, Elise Gettings^5^, Amanda Griffin^5^, Sheila Grimmel^5^, Kathleen Grinke^5^, Kathryn Hall^5^, Meg Healy^5^, Deborah Henault^5^, Grace Holland^5^, Chantal Kayitesi^5^, Vlasta LaValle^5^, Yuting Lu^5^, Sarah Luthern^5^, Jordan Marchewka (Schneider)^5^, Brittani Martino^5^, Roseann McNamara^5^, Christian Nambu^5^, Susan Nelson^5^, Marjorie Noone^5^, Christine Ommerborn^5^, Lois Chris Pacheco^5^, Nicole Phan^5^, Falisha A. Porto^5^, Edward Ryan^5^, Kathleen Selleck^5^, Sue Slaughenhaupt^5^, Kimberly Smith Sheppard^5^, Elizabeth Suschana^5^, Vivine Wilson^5^, Galit Alter^6^, Alejandro Balazs^6^, Julia Bals^6^, Max Barbash^6^, Yannic Bartsch^6^, Julie Boucau^6^, Josh Chevalier^6^, Fatema Chowdhury^6^, Kevin Einkauf^6^, Jon Fallon^6^, Liz Fedirko^6^, Kelsey Finn^6^, Pilar Garcia-Broncano^6^, Ciputra Hartana^6^, Chenyang Jiang^6^, Paulina Kaplonek^6^, Marshall Karpell^6^, Evan C. Lam^6^, Kristina Lefteri^6^, Xiaodong Lian^6^, Mathias Lichterfeld^6^, Daniel Lingwood^6^, Hang Liu^6^, Jinqing Liu^6^, Natasha Ly^6^, Ashlin Michell^6^, Ilan Millstrom^6^, Noah Miranda^6^, Claire O’Callaghan^6^, Matthew Osborn^6^, Shiv Pillai^6^, Yelizaveta Rassadkina^6^, Alexandra Reissis^6^, Francis Ruzicka^6^, Kyra Seiger^6^, Libera Sessa^6^, Christianne Sharr^6^, Sally Shin^6^, Nishant Singh^6^, Weiwei Sun^6^, Xiaoming Sun^6^, Hannah Ticheli^6^, Alicja Trocha-Piechocka^6^, Daniel Worrall^6^, Alex Zhu^6^, George Daley^7^, David Golan^7^, Howard Heller^7^, Arlene Sharpe^7^, Nikolaus Jilg^8^, Alex Rosenthal^8^, Colline Wong^8^

^1^Department of Emergency Medicine, Massachusetts General Hospital, Boston, MA, USA.

^2^Massachusetts General Hospital Cancer Center, Boston, MA, USA.

^3^Division of Infectious Diseases, Department of Medicine, Massachusetts General Hospital, Boston, MA, USA.

^4^Massachusetts General Hospital Center for Immunology and Inflammatory Diseases, Boston, MA, USA.

^5^Massachusetts General Hospital, Boston, MA, USA.

^6^Ragon Institute of MGH, MIT and Harvard, Cambridge, MA, USA.

^7^Harvard Medical School, Boston, MA, USA.

^8^Brigham and Women’s Hospital, Boston, MA, USA.

## Author information

Correspondence and requests for materials should be addressed to J.L.

