## Supplementary material for "Innate lymphoid cells and disease tolerance in SARS-CoV-2 infection": Supp Figs 1-6; Supp Tables S1-9

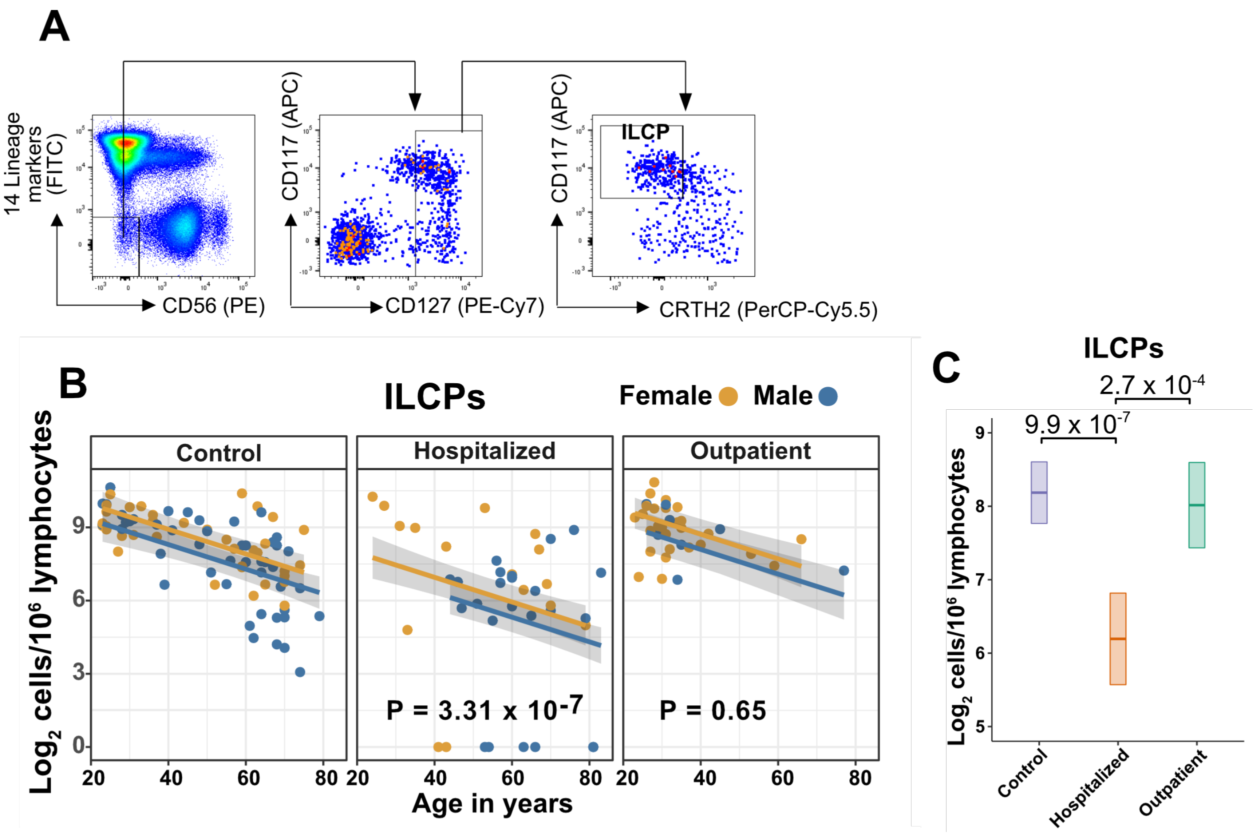


**Fig. S1. Innate lymphoid cell precursors (ILCPs) decrease with age and are depleted in patients hospitalized with COVID-19**

(A) Representative gating for ILCP identification in PBMCs. Cells were first gated on lymphoid cells, singlets, live/dead, and $CD45^+$. Lineage (Lin) markers included antibodies against: CD3, CD4, TCRαβ, TCRγδ, CD19, CD20, CD22, CD34, FcεRIα, CD11c, CD303, CD123, CD1a, and CD14.

(B) Effect of age (X-axis) on $\log_2$ ILCP abundance per million total lymphocytes (Y-axis). Each dot represents an individual blood donor, with yellow for female and blue for male. Shading represents the 95%CI. P-values are from the regression analysis for comparisons to the control group.

(C) Lymphoid cell abundance by group, shown as estimated marginal means with 95%CI, generated from the multiple linear regressions in (B), and averaged across age and sex. P-values represent pairwise comparisons on the estimated marginal means, adjusted for multiple comparisons with the Tukey method.

**A**

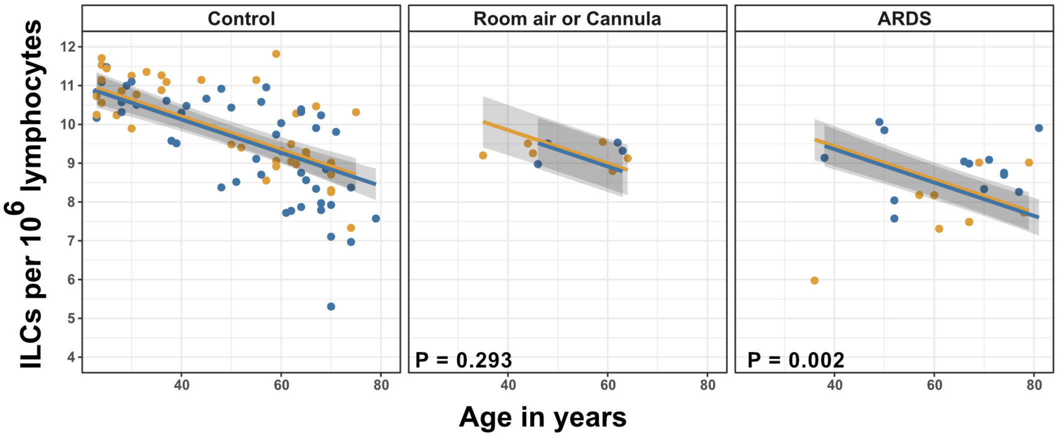


**B**

| Group differences in ILC[a] abundance adjusted for age and sex | |
|---|---|
| **Group** | **Fold difference (log2) [±95%CI]** |
| On room air or nasal cannula | -0.353 [-1.016, 0.309] |
| ARDS | -0.769** [-1.250, -0.288] |

* p < 0.05, ** p < 0.01, *** p < 0.001
[a]per $10^6$ lymphocytes

**Fig. S2. Association of blood ILC depletion with COVID-19 severity in an independent cohort of adults.**

(A) Effect of age (X-axis) on $log_2$ ILC abundance per million total lymphocytes (Y-axis) in adult controls from this paper and patients with COVID-19 from Kuri-Cervantes et al., 2020. Patients were stratified into two groups by disease severity. The first group included patients maintained on room air or treated with O2 by nasal cannula. The second group included those with ARDS. Each dot represents an individual blood donor, with yellow for female and blue for male. Shading represents the 95%CI. P-values are from the regression analysis for comparisons to the control group.

(C) Table of regression coefficients for $log_2$ fold difference in ILC abundance for patients with COVID-19, in comparison to the adult healthy control cohort, adjusted for effects of age and sex.

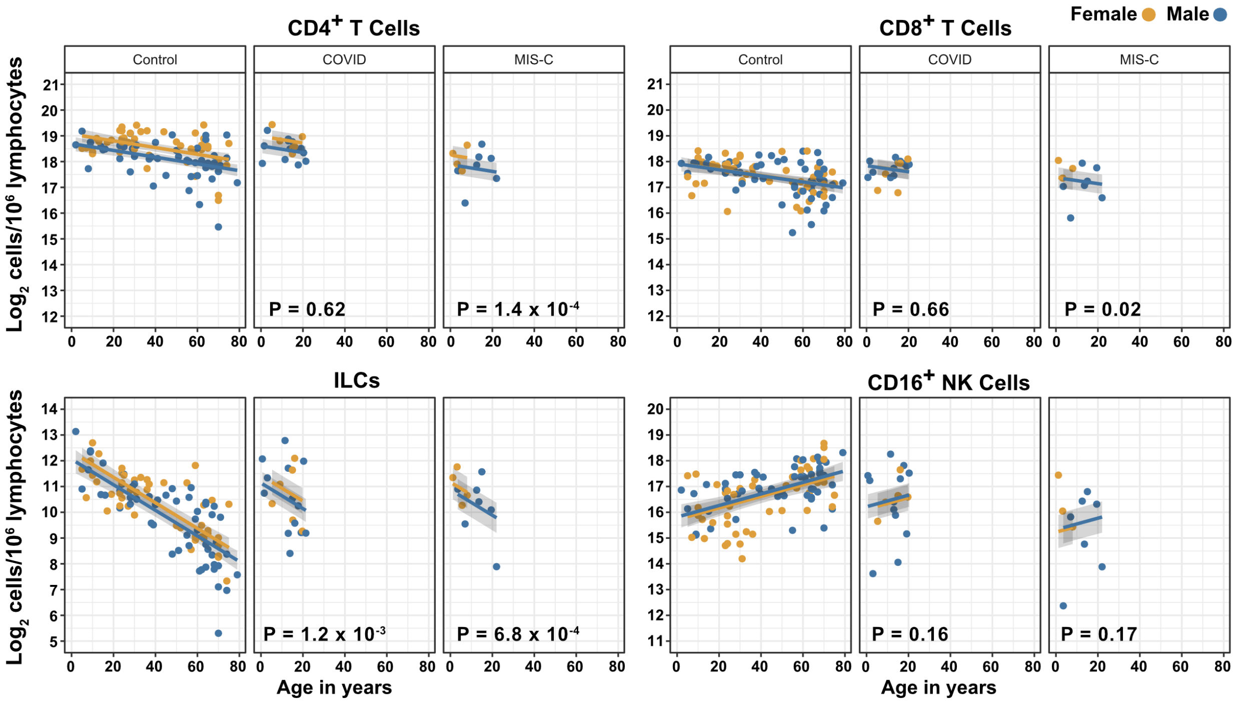


**Fig. S3. Effects of pediatric COVID-19 and MIS-C on blood lymphoid cell subsets in comparison to full combined adult and pediatric control group**

Effect of age (X-axis) on $\log_2$ abundance per million total lymphocytes of the indicated lymphoid cell populations (Y-axis). Each dot represents an individual blood donor, with yellow for female and blue for male. Shading represents the 95%CI. P-values are from the regression analysis for comparisons to the control group.

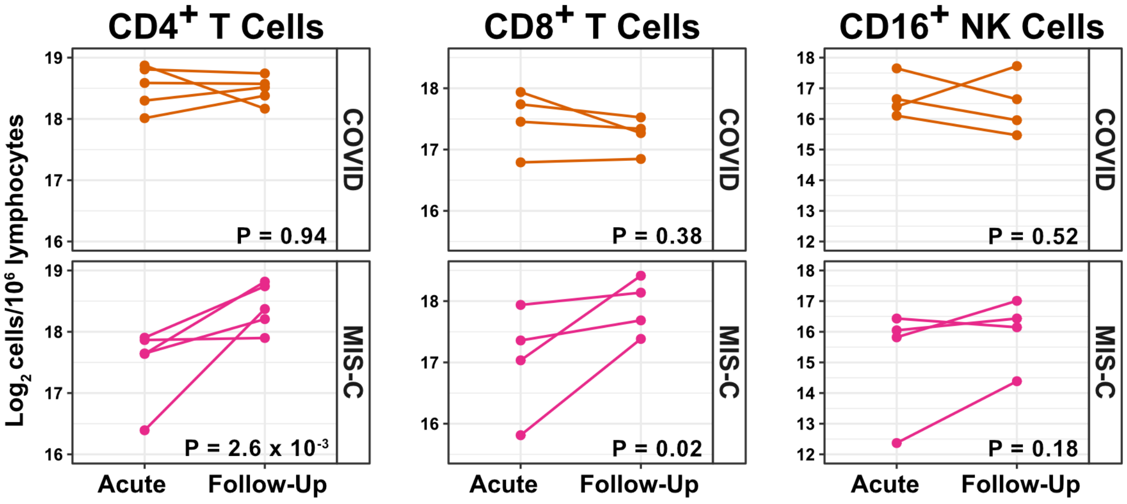


**Fig. S4. T cells increase during follow-up from MIS-C**

$Log_2$ ILC abundance per million lymphocytes in longitudinal pairs of samples collected during acute presentation and during follow-up, from individual children with COVID-19 or MIS-C. P-values are for change in ILC abundance at follow-up, as determined with a linear mixed model, adjusting for age, sex, and group, and with patient as a random effect. Differences in sample size among cell types was due to limited sample availability.

A

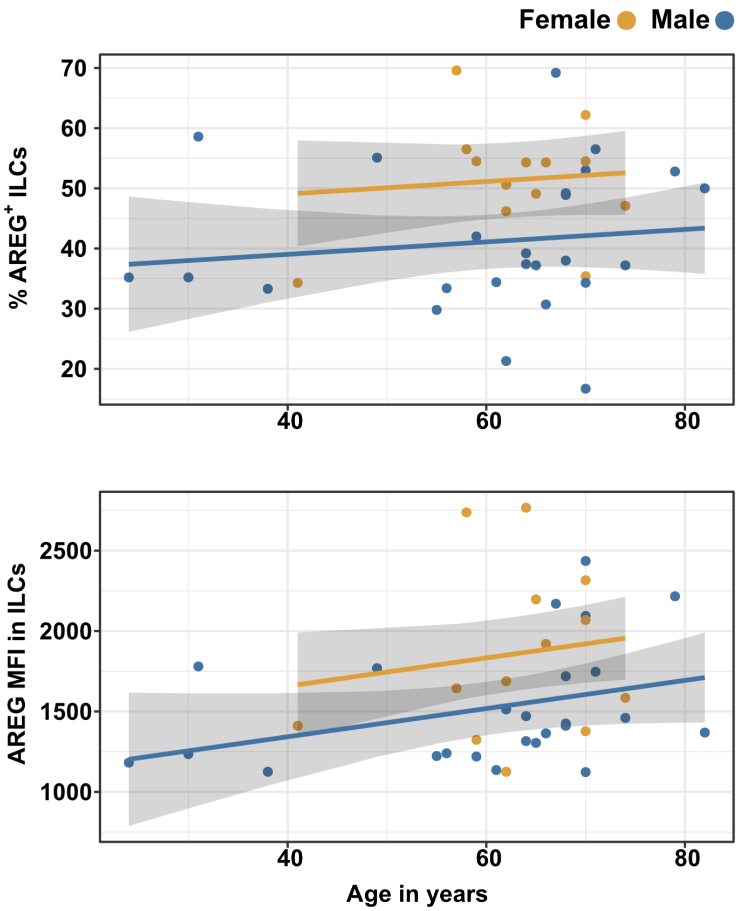


**Fig. S5. Males have lower percent AREG+ ILCs, and lower AREG MFI in ILCs, than do females, and there is no effect of age on these parameters**

(A) Effect of age (X-axis) on % AREG+ ILCs or MFI in ILCs (Y-axis as indicated). Each dot represents an individual blood donor, with yellow for female and blue for male. Shading represents the 95%CI.

(B) Table of results for regression analyses plotted in (A)

B

Difference [±95%CI]

| | **% AREG+** | **MFI in ILCs** |
|---|---|---|
| Age | 0.103 | 8.764 |
| | [-0.190, 0.397] | [-2.033, 19.561] |
| Male | -10.030* | -314.816* |
| | [-18.037, -2.023] | [-609.660, -19.973] |
| $R^2$ | 0.174 | 0.189 |

* p < 0.05, ** p < 0.01, *** p < 0.001

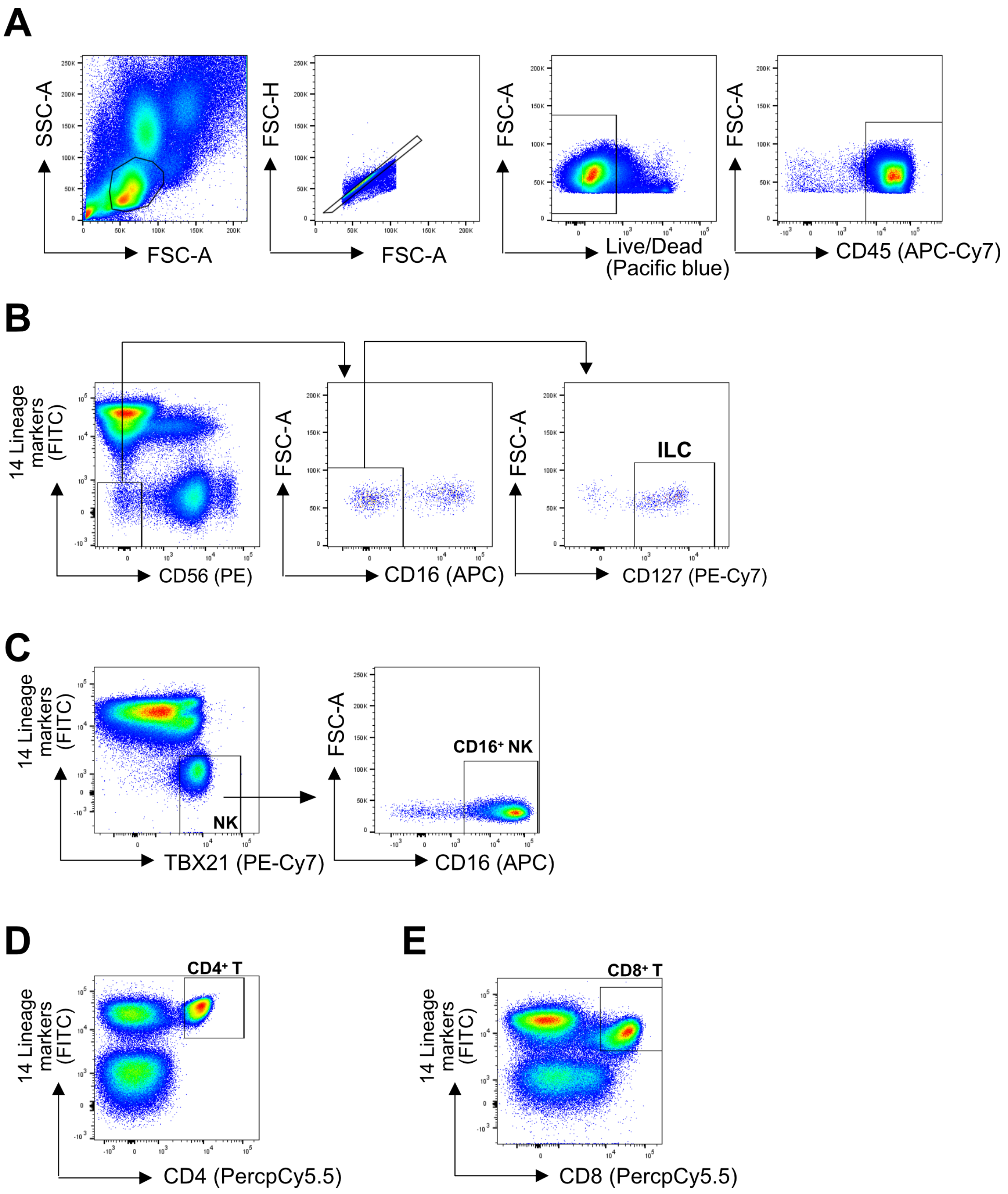


**Fig. S6. Representative gating strategy**

(A) All cell subsets were first gated on lymphoid cells, singlets, live/dead, and CD45$^{+}$. Lineage (Lin) markers included antibodies against: CD3, CD4, TCRαβ, TCRγδ, CD19, CD20, CD22, CD34, FcεRIα, CD11c, CD303, CD123, CD1a, and CD14.

(B) ILCs were identified as Lin$^{-}$CD56$^{-}$CD16$^{-}$CD127$^{+}$

(C) CD16$^{+}$ NK cells were identified as Lin$^{-}$TBX21$^{+}$CD16$^{+}$

(E) CD4$^{+}$ T cells were identified as Lin$^{+}$CD4$^{+}$ (D), and CD8$^{+}$ T cells were identified as Lin$^{+}$CD8$^{+}$

**Table S1: Race and Ethnicity of Adult Cohorts**

| Characteristic | **Healthy Control N=86** | **Hospitalized N=40** | **Outpatient N=51** |
|---|---|---|---|
| Race or ethnic group – number (%) | | | |
| White | 25 (29.1) | 17 (42.5) | 46 (90.2) |
| Black or African American | 0 (0) | 6 (15) | 2 (3.9) |
| Asian | 1 (1.2) | 1 (2.5) | 1 (2) |
| Unknown or not reported | 60 (69.8) | 16 (40) | 2 (3.9) |
| Hispanic or latinx | 1 (1.2) | 11 (27.5) | 3 (5.9) |

Percentages may not equal 100 because of rounding

| Table S2: Demographic and Clinical Characteristics of Pediatric Blood Donor Groups | | | | | |
|---|---|---|---|---|---|
| **Characteristic** | **Control N=17** | **COVID-19 N=19** | **COVID follow-up N=14** | **MIS-C N=11** | **MISC-C follow-up N=7** |
| Mean age (range) - years | 10.6 (2-19) | 13 (0.7-21.4) | 13.8 (1-22) | 10.1 (1.2-22) | 6.3 (2.2-13.1) |
| Sex – number (%) | | | | | |
| Male | 8 (47.1) | 13 (68.4) | 7 (50) | 7 (63.6) | 5 (71.4) |
| Female | 9 (52.9) | 6 (31.6) | 7 (50) | 4 (36.4) | 2 (28.6) |
| Hospital admission for COVID-19 – number (%) | | 11 (57.9) | 4 (28.6) | 0 | 0 |
| Hospital admission for MIS-C – number (%) | | | | 11 (100) | 7 (100) |
| Intubation with mechanical ventilation or ECMO – number (%) | | 1 (5.3) | 0 | 1 (9.1) | 1 (14.3) |
| Deaths – number (%) | | 0 | 0 | 0 | 0 |
| Mean time hospitalized (range) – days | | 8 (2-17) | | 7.5 (1-16) | |
| Max lab value – mean (range) | | | | | |
| CRP – mg/L | | 119 (0.2-385) | | 161 (9-300) | |

**Table S3: Change in Lymphocyte Abundance Due to Age, Sex, and COVID-19 Severity in Adult cohorts**

Fold difference (log2) [±95%CI]

| | Lymphocytes[a] |
|---|---|
| Age | -0.008*** [-0.012, -0.004] |
| Male | -0.267*** [-0.400, -0.135] |
| Hospitalized | -0.411*** [-0.572, -0.250] |
| Outpatient | 0.129 [-0.030, 0.289] |
| $R^2$ | 0.416 |

* p < 0.05, ** p < 0.01, *** p < 0.001

[a]per $10^6$ PBMCs ‘

**Table S4: Change in Cell Abundance Due to Age, Sex, and COVID-19 Severity**

Fold difference (log2) [±95%CI]

| | **$CD4^+$ T[a]** | **ILC[a]** | **$CD8^+$ T[a]** | **$CD16^+$ NK[a]** |
|---|---|---|---|---|
| Age | -0.020*** | -0.051*** | -0.017*** | 0.014* |
| | [-0.029, -0.010] | [-0.063, -0.040] | [-0.025, -0.008] | [0.002, 0.026] |
| Male | -0.678*** | -0.609** | -0.411** | -0.049 |
| | [-0.980, -0.375] | [-0.983, -0.235] | [-0.688, -0.134] | [-0.435, 0.336] |
| Hospitalized | -0.241 | -1.280*** | -0.034 | -1.466*** |
| | [-0.608, 0.126] | [-1.734, -0.826] | [-0.369, 0.301] | [-1.932, -0.999] |
| Outpatient | 0.461* | 0.009 | 0.224 | -0.276 |
| | [0.099, 0.824] | [-0.441, 0.458] | [-0.110, 0.558] | [-0.741, 0.189] |
| $R^2$ | 0.364 | 0.552 | 0.224 | 0.194 |

* p < 0.05, ** p < 0.01, *** p < 0.001
[a]per $10^6$ PBMCs

**Table S5: Odds of Hospitalization in Pediatric Cohort[a]**

| Cell count[b] | Odds Ratio[c] | 95% Confidence Interval | P-Value |
|---|---|---|---|
| **$CD4^+$ T** | 1.75 | 0.0593 – 43.3 | 0.724 |
| **ILC** | 0.712 | 0.26– 1.73 | 0.463 |
| **$CD8^+$ T** | 0.217 | 0.00509 – 5.9 | 0.366 |
| **$CD16^+$ NK** | 0.386 | 0.0554 – 1.23 | 0.194 |

[a]Adjusted for age and sex
[b]per $10^6$ lymphocytes
[c]per 2-fold increase in cell population abundance

**Table S6: Change in Pediatric Cohort Cell Abundance due to Group; Adjusted for Effects of Age and Sex with Combined Pediatric and Adult Control Data**

Fold difference (log2) [±95%CI]

| | $CD4^+$ T[a] | ILC[a] | $CD8^+$ T[a] | $CD16^+$ NK[a] |
|---|---|---|---|---|
| COVID | -0.085 | -0.924** | -0.078 | 0.400 |
| | [-0.421, 0.251] | [-1.474, -0.373] | [-0.424, 0.269] | [-0.161, 0.961] |
| MIS-C | -0.814*** | -1.183*** | -0.538* | -0.493 |
| | [-1.224, -0.404] | [-1.856, -0.511] | [-0.969, -0.106] | [-1.190, 0.205] |
| $R^2$ | 0.292 | 0.541 | 0.165 | 0.261 |

* $p < 0.05$, ** $p < 0.01$, *** $p < 0.001$
[a]per $10^6$ lymphocytes

| Table S7: Gene ontology analysis results | | | | | | | | |
|---|---|---|---|---|---|---|---|---|
| ID | Description | GeneRatio | BgRatio | pvalue | p.adjust | qvalue | geneID | Count |
| GO:0032616 | interleukin-13 production | 7/294 | 17/9263 | 4.47E-07 | 0.00112026 | 0.00100638 | IL17RA/NLRP3/IL18/TNFSF4/ARG2/GATA3/RARA | 7 |
| GO:0042092 | type 2 immune response | 8/294 | 25/9263 | 6.33E-07 | 0.00112026 | 0.00100638 | NLRP3/CD81/IL18/BCL3/TNFSF4/ARG2/GATA3/RARA | 8 |
| GO:0002711 | positive regulation of T cell mediated immunity | 9/294 | 36/9263 | 1.28E-06 | 0.00122325 | 0.0010989 | NLRP3/CD81/IL18/HLA-B/IL23R/HSPD1/TNFSF4/HLA-A/GATA3 | 9 |
| GO:0045064 | T-helper 2 cell differentiation | 6/294 | 13/9263 | 1.38E-06 | 0.00122325 | 0.0010989 | NLRP3/IL18/BCL3/TNFSF4/GATA3/RARA | 6 |
| GO:0002830 | positive regulation of type 2 immune response | 6/294 | 14/9263 | 2.35E-06 | 0.00123526 | 0.00110969 | NLRP3/CD81/IL18/TNFSF4/GATA3/RARA | 6 |
| GO:0002828 | regulation of type 2 immune response | 7/294 | 21/9263 | 2.40E-06 | 0.00123526 | 0.00110969 | NLRP3/CD81/IL18/TNFSF4/ARG2/GATA3/RARA | 7 |
| GO:0002709 | regulation of T cell mediated immunity | 10/294 | 49/9263 | 2.44E-06 | 0.00123526 | 0.00110969 | NLRP3/CD81/IL18/HLA-B/IL23R/HSPD1/TNFSF4/HLA-A/GATA3/WAS | 10 |
| GO:0032656 | regulation of interleukin-13 production | 6/294 | 15/9263 | 3.82E-06 | 0.00169036 | 0.00151854 | IL17RA/NLRP3/TNFSF4/ARG2/GATA3/RARA | 6 |
| GO:0002708 | positive regulation of lymphocyte mediated immunity | 12/294 | 79/9263 | 6.38E-06 | 0.00212609 | 0.00190997 | NLRP3/CD81/IL18/HLA-B/CD28/LAMP1/IL23R/HSPD1/TNFSF4/HLA-A/GATA3/EXOSC6 | 12 |
| GO:0002706 | regulation of lymphocyte mediated immunity | 14/294 | 107/9263 | 6.60E-06 | 0.00212609 | 0.00190997 | NLRP3/CD81/IL18/HLA-B/CD28/LAMP1/IL23R/HSPD1/TNFSF4/HLA-A/ARRB2/GATA3/EXOSC6/WAS | 14 |
| GO:0046631 | alpha-beta T cell activation | 14/294 | 107/9263 | 6.60E-06 | 0.00212609 | 0.00190997 | RASAL3/FOXP1/NLRP3/CD81/IL18/RC3H1/CD28/BCL3/IL23R/TCF7/TNFSF4/ARG2/GATA3/RARA | 14 |
| GO:0035710 | CD4-positive, alpha-beta T cell activation | 11/294 | 69/9263 | 9.66E-06 | 0.00274122 | 0.00246257 | FOXP1/NLRP3/CD81/IL18/RC3H1/BCL3/IL23R/TNFSF4/ARG2/GATA3/RARA | 11 |

| | | | | | | | | |
|---|---|---|---|---|---|---|---|---|
| GO:0051251 | positive regulation of lymphocyte activation | 20/294 | 209/9263 | 1.02E-05 | 0.00274122 | 0.00246257 | CD6/RASAL3/NLRP3/GRAP2/CD81/IL18/CD28/FLT3LG/CD5/LAMP1/IL23R/HSPD1/TNFSF4/RAC1/CDC42/ADAM8/CSK/GATA3/RARA/EXOSC6 | 20 |
| GO:0050870 | positive regulation of T cell activation | 17/294 | 159/9263 | 1.11E-05 | 0.00274122 | 0.00246257 | CD6/RASAL3/NLRP3/GRAP2/CD81/IL18/CD28/CD5/IL23R/HSPD1/TNFSF4/RAC1/CDC42/ADAM8/CSK/GATA3/RARA | 17 |
| GO:0032736 | positive regulation of interleukin-13 production | 5/294 | 23316 | 1.23E-05 | 0.00274122 | 0.00246257 | IL17RA/NLRP3/TNFSF4/GATA3/RARA | 5 |
| GO:0042093 | T-helper cell differentiation | 9/294 | 47/9263 | 1.36E-05 | 0.00274122 | 0.00246257 | FOXP1/NLRP3/IL18/RC3H1/BCL3/IL23R/TNFSF4/GATA3/RARA | 9 |
| GO:2000514 | regulation of CD4-positive, alpha-beta T cell activation | 9/294 | 47/9263 | 1.36E-05 | 0.00274122 | 0.00246257 | NLRP3/CD81/IL18/RC3H1/IL23R/TNFSF4/ARG2/GATA3/RARA | 9 |
| GO:0050863 | regulation of T cell activation | 21/294 | 232/9263 | 1.44E-05 | 0.00274122 | 0.00246257 | CD6/RASAL3/CASP3/NLRP3/GRAP2/CD81/IL18/RC3H1/CD28/CD5/IL23R/HSPD1/LAT/TNFSF4/RAC1/ARG2/CDC42/ADAM8/CSK/GATA3/RARA | 21 |
| GO:0046634 | regulation of alpha-beta T cell activation | 11/294 | 72/9263 | 1.47E-05 | 0.00274122 | 0.00246257 | RASAL3/NLRP3/CD81/IL18/RC3H1/CD28/IL23R/TNFSF4/ARG2/GATA3/RARA | 11 |
| GO:0046649 | lymphocyte activation | 34/294 | 498/9263 | 1.83E-05 | 0.00302412 | 0.00271671 | CD6/RASAL3/FOXP1/CASP3/NLRP3/GRAP2/CD81/IL18/RC3H1/IRF2BP2/CD28/FLT3LG/CD5/HDAC9/LAMP1/BCL3/PLCG2/IL23R/AKAP17A/TCF7/HSPD1/LAT/TNFSF4/RAC1/ARG2/ZBTB7A/CDC42/ADAM8/CSK/GATA3/RARA/EXOSC6/STK11/WAS | 34 |
| GO:0002824 | positive regulation of | 11/294 | 74/9263 | 1.92E-05 | 0.00302412 | 0.00271671 | NLRP3/CD81/IL18/HLA- | 11 |

| | | | | | | | | |
|---|---|---|---|---|---|---|---|---|
| | adaptive immune response based on somatic recombination of immune receptors built from immunoglobulin superfamily domains | | | | | | B/CD28/IL23R/HSPD1/TNFSF4/HLA-A/GATA3/EXOSC6 | |
| GO:0002294 | CD4-positive, alpha-beta T cell differentiation involved in immune response | 9/294 | 49/9263 | 1.94E-05 | 0.00302412 | 0.00271671 | FOXP1/NLRP3/IL18/RC3H1/BCL3/IL23R/TNFSF4/GATA3/RARA | 9 |
| GO:0002822 | regulation of adaptive immune response based on somatic recombination of immune receptors built from immunoglobulin superfamily domains | 13/294 | 103/9263 | 2.10E-05 | 0.00302412 | 0.00271671 | NLRP3/CD81/IL18/RC3H1/HLA-B/CD28/IL23R/HSPD1/TNFSF4/HLA-A/GATA3/EXOSC6/WAS | 13 |
| GO:0022407 | regulation of cell-cell adhesion | 23/294 | 276/9263 | 2.16E-05 | 0.00302412 | 0.00271671 | CD6/RASAL3/CASP3/NLRP3/GRAP2/CD81/IL18/RC3H1/CD28/CD5/TNR/IL23R/ADAM19/HSPD1/TNFSF4/RAC1/ARG2/CDC42/ADAM8/CSK/GATA3/RGCC/RARA | 23 |
| GO:0022409 | positive regulation of cell-cell adhesion | 18/294 | 185/9263 | 2.27E-05 | 0.00302412 | 0.00271671 | CD6/RASAL3/NLRP3/GRAP2/CD81/IL18/CD28/CD5/IL23R/ADAM19/HSPD1/TNFSF4/RAC1/CDC42/ADAM8/CSK/GATA3/RARA | 18 |
| GO:0002287 | alpha-beta T cell activation involved in immune response | 9/294 | 50/9263 | 2.31E-05 | 0.00302412 | 0.00271671 | FOXP1/NLRP3/IL18/RC3H1/BCL3/IL23R/TNFSF4/GATA3/RARA | 9 |
| GO:0002293 | alpha-beta T cell differentiatio | 9/294 | 50/9263 | 2.31E-05 | 0.00302412 | 0.00271671 | FOXP1/NLRP3/IL18/RC3H1/BCL3/I | 9 |

| | | | | | | | | |
|---|---|---|---|---|---|---|---|---|
| | n involved in immune response | | | | | | L23R/TNFSF4/GATA3/RARA | |
| GO:0002285 | lymphocyte activation involved in immune response | 15/294 | 136/9263 | 2.54E-05 | 0.00321286 | 0.00288626 | FOXP1/NLRP3/CD81/IL18/RC3H1/CD28/LAMP1/BCL3/PLCG2/IL23R/HSPD1/TNFSF4/GATA3/RARA/EXOSC6 | 15 |
| GO:1903039 | positive regulation of leukocyte cell-cell adhesion | 17/294 | 171/9263 | 2.90E-05 | 0.00354228 | 0.0031822 | CD6/RASAL3/NLRP3/GRAP2/CD81/IL18/CD28/CD5/IL23R/HSPD1/TNFSF4/RAC1/CDC42/ADAM8/CSK/GATA3/RARA | 17 |
| GO:1903037 | regulation of leukocyte cell-cell adhesion | 20/294 | 225/9263 | 3.01E-05 | 0.00354723 | 0.00318665 | CD6/RASAL3/CASP3/NLRP3/GRAP2/CD81/IL18/RC3H1/CD28/CD5/IL23R/HSPD1/TNFSF4/RAC1/ARG2/CDC42/ADAM8/CSK/GATA3/RARA | 20 |
| GO:0042110 | T cell activation | 26/294 | 344/9263 | 3.46E-05 | 0.00384309 | 0.00345243 | CD6/RASAL3/FOXP1/CASP3/NLRP3/GRAP2/CD81/IL18/RC3H1/CD28/CD5/BCL3/IL23R/TCF7/HSPD1/LAT/TNFSF4/RAC1/ARG2/CDC42/ADAM8/CSK/GATA3/RARA/STK11/WAS | 26 |
| GO:0045785 | positive regulation of cell adhesion | 22/294 | 265/9263 | 3.47E-05 | 0.00384309 | 0.00345243 | CD6/RASAL3/NLRP3/GRAP2/CD81/IL18/DOCK5/CD28/CD5/SPOCK2/IL23R/ADAM19/HSPD1/RSU1/TNFSF4/RAC1/CDC42/ADAM8/CSK/GATA3/RARA/ITGB1BP1 | 22 |
| GO:0002821 | positive regulation of adaptive immune response | 11/294 | 79/9263 | 3.61E-05 | 0.00387396 | 0.00348017 | NLRP3/CD81/IL18/HLA-B/CD28/IL23R/HSPD1/TNFSF4/HLA-A/GATA3/EXOSC6 | 11 |
| GO:0046640 | regulation of alpha-beta T cell proliferation | 6/294 | 22/9263 | 4.72E-05 | 0.00482039 | 0.00433039 | RASAL3/CD81/IL18/CD28/TNFSF4/ARG2 | 6 |
| GO:0051249 | regulation of lymphocyte activation | 24/294 | 310/9263 | 4.76E-05 | 0.00482039 | 0.00433039 | CD6/RASAL3/CASP3/NLRP3/GRAP2/CD81/IL18/RC3H1/CD28/FLT3LG/CD5/LAMP1/IL | 24 |

| | | | | | | | | |
|---|---|---|---|---|---|---|---|---|
| | | | | | | | 23R/HSPD1/LAT/TNFSF4/RAC1/ARG2/CDC42/ADAM8/CSK/GATA3/RARA/EXOSC6 | |
| GO:0002696 | positive regulation of leukocyte activation | 20/294 | 235/9263 | 5.59E-05 | 0.00549879 | 0.00493983 | CD6/RASAL3/NLRP3/GRAP2/CD81/IL18/CD28/FLT3LG/CD5/LAMP1/IL23R/HSPD1/TNFSF4/RAC1/CDC42/ADAM8/CSK/GATA3/RARA/EXOSC6 | 20 |
| GO:0002292 | T cell differentiation involved in immune response | 9/294 | 56/9263 | 5.90E-05 | 0.00564657 | 0.00507259 | FOXP1/NLRP3/IL18/RC3H1/BCL3/IL23R/TNFSF4/GATA3/RARA | 9 |
| GO:0043367 | CD4-positive, alpha-beta T cell differentiation | 9/294 | 57/9263 | 6.81E-05 | 0.00634978 | 0.00570432 | FOXP1/NLRP3/IL18/RC3H1/BCL3/IL23R/TNFSF4/GATA3/RARA | 9 |
| GO:0050865 | regulation of cell activation | 27/294 | 381/9263 | 7.46E-05 | 0.00677255 | 0.00608411 | CD6/RASAL3/CASP3/NLRP3/GRAP2/CD81/IL18/RC3H1/CD28/FLT3LG/CD5/LAMP1/LMO4/IL23R/HSPD1/TSPAN32/LAT/TNFSF4/RAC1/ARG2/CDC42/ADAM8/CSK/GATA3/RARA/EXOSC6/TEC | 27 |
| GO:0001817 | regulation of cytokine production | 31/294 | 468/9263 | 7.76E-05 | 0.00686707 | 0.00616902 | PCBP2/HSF1/CD6/FOXP1/HSPA1B/IL17RA/NLRP3/CD81/IL18/IRAK3/DDX3X/LTB/CD28/HDAC9/HSPA1A/BCL3/TXK/PLCG2/IFNGR1/IL23R/HSPD1/TMSB4X/TNFSF4/RAC1/ARG2/ADAM8/CSK/ARRB2/GATA3/RGCC/RARA | 31 |
| GO:0050867 | positive regulation of cell activation | 20/294 | 241/9263 | 7.96E-05 | 0.00687411 | 0.00617535 | CD6/RASAL3/NLRP3/GRAP2/CD81/IL18/CD28/FLT3LG/CD5/LAMP1/IL23R/HSPD1/TNFSF4/RAC1/CDC42/ADAM8/CSK/GATA3/RARA/EXOSC6 | 20 |
| GO:0031295 | T cell costimulation | 8/294 | 46/9263 | 8.54E-05 | 0.00701177 | 0.00629902 | GRAP2/CD81/CD28/CD5/TNFSF4/ | 8 |

| | | | | | | | | |
|---|---|---|---|---|---|---|---|---|
| | | | | | | | RAC1/CDC42/CSK | |
| GO:0030155 | regulation of cell adhesion | 29/294 | 427/9263 | 8.60E-05 | 0.00701177 | 0.00629902 | CD6/RASAL3/CASP3/CD164/NLRP3/GRAP2/CD81/IL18/RC3H1/DOCK5/CD28/CD5/SPOCK2/TNR/IL23R/ADAM19/HSPD1/RSU1/TNFSF4/RAC1/ARG2/CDC42/ADAM8/CSK/ARHGDIA/GATA3/RGCC/RARA/ITGB1BP1 | 29 |
| GO:0002705 | positive regulation of leukocyte mediated immunity | 12/294 | 102/9263 | 8.78E-05 | 0.00701177 | 0.00629902 | NLRP3/CD81/IL18/HLA-B/CD28/LAMP1/IL23R/HSPD1/TNFSF4/HLA-A/GATA3/EXOSC6 | 12 |
| GO:0002819 | regulation of adaptive immune response | 13/294 | 118/9263 | 8.91E-05 | 0.00701177 | 0.00629902 | NLRP3/CD81/IL18/RC3H1/HLA-B/CD28/IL23R/HSPD1/TNFSF4/HLA-A/GATA3/EXOSC6/WAS | 13 |
| GO:0031294 | lymphocyte costimulation | 8/294 | 47/9263 | 0.00010016 | 0.00771021 | 0.00692646 | GRAP2/CD81/CD28/CD5/TNFSF4/RAC1/CDC42/CSK | 8 |
| GO:0046633 | alpha-beta T cell proliferation | 6/294 | 25/9263 | 0.00010332 | 0.00778396 | 0.00699271 | RASAL3/CD81/IL18/CD28/TNFSF4/ARG2 | 6 |
| GO:0007159 | leukocyte cell-cell adhesion | 20/294 | 247/9263 | 0.00011181 | 0.0082481 | 0.00740967 | CD6/RASAL3/CASP3/NLRP3/GRAP2/CD81/IL18/RC3H1/CD28/CD5/IL23R/HSPD1/TNFSF4/RAC1/ARG2/CDC42/ADAM8/CSK/GATA3/RARA | 20 |
| GO:0045622 | regulation of T-helper cell differentiation | 6/294 | 26/9263 | 0.00013077 | 0.00922661 | 0.00828871 | NLRP3/IL18/RC3H1/IL23R/TNFSF4/RARA | 6 |
| GO:0002286 | T cell activation involved in immune response | 10/294 | 76/9263 | 0.00013302 | 0.00922661 | 0.00828871 | FOXP1/NLRP3/CD81/IL18/RC3H1/BCL3/IL23R/TNFSF4/GATA3/RARA | 10 |
| GO:0043370 | regulation of CD4-positive, alpha-beta T cell differentiation | 7/294 | 37/9263 | 0.00013713 | 0.00922661 | 0.00828871 | NLRP3/IL18/RC3H1/IL23R/TNFSF4/GATA3/RARA | 7 |
| GO:0002726 | positive regulation of | 5/294 | 17/9263 | 0.00014071 | 0.00922661 | 0.00828871 | NLRP3/CD81/IL18/TNFSF4/GATA3 | 5 |

| | | | | | | | | |
|---|---|---|---|---|---|---|---|---|
| | T cell cytokine production | | | | | | | |
| GO:00 45624 | positive regulation of T-helper cell differentiatio n | 5/294 | 17/9263 | 0.0001 4071 | 0.009226 61 | 0.00828 871 | NLRP3/IL18/IL23 R/TNFSF4/RARA | 5 |
| GO:00 46641 | positive regulation of alpha-beta T cell proliferation | 5/294 | 17/9263 | 0.0001 4071 | 0.009226 61 | 0.00828 871 | RASAL3/CD81/IL 18/CD28/TNFSF4 | 5 |
| GO:00 97190 | apoptotic signaling pathway | 29/294 | 440/926 3 | 0.0001 4545 | 0.009364 48 | 0.00841 257 | YWHAZ/TRADD/ BCL2L12/CASP3/ HSPA1B/GNAI2/X PA/HINT1/DDX3X /MPV17L/MAL/TN FSF12/CD28/RR M2B/CD5/CHCHD 10/LAMP1/HSPA1 A/SLC9A3R1/BCL 3/TMC8/YWHAB/ SKIL/ARRB2/JUN /PPP3CC/STK11/ CDKN2D/IFI6 | 29 |
| GO:00 02456 | T cell mediated immunity | 10/294 | 77/9263 | 0.0001 4866 | 0.009399 95 | 0.00844 443 | NLRP3/CD81/IL1 8/HLA- B/IL23R/HSPD1/T NFSF4/HLA- A/GATA3/WAS | 10 |
| GO:00 46635 | positive regulation of alpha-beta T cell activation | 8/294 | 50/9263 | 0.0001 5745 | 0.009781 49 | 0.00878 719 | RASAL3/NLRP3/ CD81/IL18/CD28/I L23R/TNFSF4/RA RA | 8 |
| GO:00 32754 | positive regulation of interleukin-5 production | 4/294 | 10/9263 | 0.0001 7948 | 0.010772 11 | 0.00967 711 | IL17RA/NLRP3/G ATA3/RARA | 4 |
| GO:00 45628 | regulation of T-helper 2 cell differentiatio n | 4/294 | 10/9263 | 0.0001 7948 | 0.010772 11 | 0.00967 711 | NLRP3/IL18/TNF SF4/RARA | 4 |
| GO:00 32753 | positive regulation of interleukin-4 production | 5/294 | 18/9263 | 0.0001 898 | 0.011201 17 | 0.01006 255 | NLRP3/CD28/TN FSF4/GATA3/RA RA | 5 |
| GO:00 02694 | regulation of leukocyte activation | 25/294 | 361/926 3 | 0.0001 9874 | 0.011536 73 | 0.01036 401 | CD6/RASAL3/CA SP3/NLRP3/GRA P2/CD81/IL18/RC 3H1/CD28/FLT3L G/CD5/LAMP1/IL 23R/HSPD1/TSP AN32/LAT/TNFSF 4/RAC1/ARG2/CD C42/ADAM8/CSK/ GATA3/RARA/EX OSC6 | 25 |
| GO:00 46632 | alpha-beta T cell | 10/294 | 80/9263 | 0.0002 0518 | 0.011611 93 | 0.01043 156 | FOXP1/NLRP3/IL 18/RC3H1/BCL3/I | 10 |

| | | | | | | | | |
|---|---|---|---|---|---|---|---|---|
| | differentiatio n | | | | | | L23R/TCF7/TNFS F4/GATA3/RARA | |
| GO:00 02250 | adaptive immune response | 22/294 | 299/926 3 | 0.0002 066 | 0.011611 93 | 0.01043 156 | CD6/NLRP3/CD8 1/IL18/RC3H1/LI ME1/HLA-B/TAP2/CD28/BC L3/TXK/IL23R/HS PD1/LAT/TNFSF4 /ARG2/CSK/HLA-A/GATA3/EXOSC 6/TEC/WAS | 22 |
| GO:00 01818 | negative regulation of cytokine production | 16/294 | 182/926 3 | 0.0002 1414 | 0.011847 82 | 0.01064 347 | PCBP2/HSF1/NL RP3/IRAK3/HDAC 9/BCL3/IL23R/TM SB4X/TNFSF4/R AC1/ARG2/CSK/A RRB2/GATA3/RG CC/RARA | 16 |
| GO:20 00516 | positive regulation of CD4-positive, alpha-beta T cell activation | 6/294 | 29/9263 | 0.0002 4901 | 0.013483 73 | 0.01211 309 | NLRP3/CD81/IL1 8/IL23R/TNFSF4/ RARA | 6 |
| GO:00 98609 | cell-cell adhesion | 27/294 | 410/926 3 | 0.0002 5132 | 0.013483 73 | 0.01211 309 | CD6/RASAL3/CA SP3/CD164/NLRP 3/GRAP2/CD81/IL 18/RC3H1/CD28/ CD5/TNR/IL23R/A DAM19/HSPD1/T SPAN32/PCDH9/ TNFSF4/RAC1/A RG2/CDC42/ADA M8/CSK/GATA3/ RGCC/RARA/BL OC1S4 | 27 |
| GO:19 03311 | regulation of mRNA metabolic process | 20/294 | 266/926 3 | 0.0003 0239 | 0.015981 28 | 0.01435 676 | YWHAZ/HSF1/NU P98/HNRNPA0/R C3H1/YBX1/ELAV L1/HSPA1A/SFS WAP/SNRNP70/R BM17/SF3B4/AN P32A/U2AF2/CPS F7/YWHAB/KHSR P/ZBTB7A/PTBP1 /EXOSC6 | 20 |
| GO:00 07264 | small GTPase mediated signal transduction | 24/294 | 350/926 3 | 0.0003 0894 | 0.016087 76 | 0.01445 242 | TNK2/RASAL3/G RAP2/SH3BP1/M APKAP1/ABCA1/ DOCK5/CFL1/SP RY1/STMN3/PSD 4/DNMT1/GDI1/R AB11B/RAB2A/LA T/PLEKHG3/RAC 1/TAGAP/CDC42/ ARHGEF18/ARH GDIA/JUN/WAS | 24 |
| GO:00 45197 | establishme nt or maintenance of epithelial | 5/294 | 20/9263 | 0.0003 2597 | 0.016597 48 | 0.01491 032 | ANK1/SH3BP1/SL C9A3R1/MARK2/ CDC42 | 5 |

| | | | | | | | | |
|---|---|---|---|---|---|---|---|---|
| | cell apical/basal polarity | | | | | | | |
| GO:0002703 | regulation of leukocyte mediated immunity | 14/294 | 152/9263 | 0.00032811 | 0.01659748 | 0.01491032 | NLRP3/CD81/IL18/HLA-B/CD28/LAMP1/IL23R/HSPD1/TNFSF4/HLA-A/ARRB2/GATA3/EXOSC6/WAS | 14 |
| GO:0051645 | Golgi localization | 4/294 | 12/9263 | 0.00040227 | 0.01966457 | 0.01766564 | YWHAZ/SLC9A3R1/CDC42/STK11 | 4 |
| GO:0002724 | regulation of T cell cytokine production | 5/294 | 21/9263 | 0.00041682 | 0.01966457 | 0.01766564 | NLRP3/CD81/IL18/TNFSF4/GATA3 | 5 |
| GO:0032673 | regulation of interleukin-4 production | 5/294 | 21/9263 | 0.00041682 | 0.01966457 | 0.01766564 | NLRP3/CD28/TNFSF4/GATA3/RARA | 5 |
| GO:0035088 | establishment or maintenance of apical/basal cell polarity | 5/294 | 21/9263 | 0.00041682 | 0.01966457 | 0.01766564 | ANK1/SH3BP1/SLC9A3R1/MARK2/CDC42 | 5 |
| GO:0061245 | establishment or maintenance of bipolar cell polarity | 5/294 | 21/9263 | 0.00041682 | 0.01966457 | 0.01766564 | ANK1/SH3BP1/SLC9A3R1/MARK2/CDC42 | 5 |
| GO:0032655 | regulation of interleukin-12 production | 7/294 | 44/9263 | 0.00042206 | 0.01966457 | 0.01766564 | FOXP1/IRAK3/LTB/IL23R/HSPD1/TNFSF4/ARRB2 | 7 |
| GO:0032615 | interleukin-12 production | 7/294 | 45/9263 | 0.00048644 | 0.02237001 | 0.02009606 | FOXP1/IRAK3/LTB/IL23R/HSPD1/TNFSF4/ARRB2 | 7 |
| GO:1902532 | negative regulation of intracellular signal transduction | 23/294 | 340/9263 | 0.00049896 | 0.02265149 | 0.02034894 | BCL2L12/RASAL3/GNAI2/SESN1/NLRP3/IRAK3/DDX3X/SH3BP1/MAPKAP1/RRM2B/CRTC3/SPRY1/TAOK3/HSPA1A/STMN3/SLC9A3R1/TMSB4X/CSK/ARRB2/STK11/ITGB1BP1/CDKN2D/CALM3 | 23 |
| GO:0008285 | negative regulation of cell population proliferation | 25/294 | 384/9263 | 0.00050608 | 0.02268384 | 0.020378 | HSF1/MXD4/CASP3/HSPA1B/CD164/RC3H1/SPRY1/HSPA1A/SLC9A3R1/KLF13/TSPAN32/HPGDS/ARG2/CSK/DACH1/JUN/GATA3/SKI/RGCC/RARA/SF1/STK11/ITGB1BP1/CDKN2D/PTGDR2 | 25 |

| | | | | | | | | |
|---|---|---|---|---|---|---|---|---|
| GO:0050778 | positive regulation of immune response | 29/294 | 475/9263 | 0.00052503 | 0.02323912 | 0.02087683 | FOXP1/NLRP3/GRAP2/CD81/IL18/RC3H1/LIME1/HLA-B/KRT1/CD28/LAMP1/TXK/PLCG2/IL23R/HSPD1/LAT/TNFSF4/RAC1/CDC42/ADAM8/CSK/HLA-A/GATA3/RGCC/RARA/EXOSC6/TEC/STK11/WAS | 29 |
| GO:0007265 | Ras protein signal transduction | 20/294 | 278/9263 | 0.00053512 | 0.02339329 | 0.02101533 | RASAL3/GRAP2/MAPKAP1/ABCA1/CFL1/SPRY1/STMN3/PSD4/DNMT1/GDI1/RAB11B/RAB2A/LAT/PLEKHG3/RAC1/CDC42/ARHGEF18/ARHGDIA/JUN/WAS | 20 |
| GO:0032634 | interleukin-5 production | 4/294 | 13/9263 | 0.00056661 | 0.02417319 | 0.02171595 | IL17RA/NLRP3/GATA3/RARA | 4 |
| GO:0032674 | regulation of interleukin-5 production | 4/294 | 13/9263 | 0.00056661 | 0.02417319 | 0.02171595 | IL17RA/NLRP3/GATA3/RARA | 4 |
| GO:0010631 | epithelial cell migration | 14/294 | 161/9263 | 0.00058925 | 0.02454725 | 0.02205199 | EMC10/PFN1/FOXP1/SH3BP1/TNFSF12/DOCK5/HDAC9/TMSB4X/JUN/GATA3/RGCC/ITGB1BP1/GIPC1/CORO1B | 14 |
| GO:0090132 | epithelium migration | 14/294 | 161/9263 | 0.00058925 | 0.02454725 | 0.02205199 | EMC10/PFN1/FOXP1/SH3BP1/TNFSF12/DOCK5/HDAC9/TMSB4X/JUN/GATA3/RGCC/ITGB1BP1/GIPC1/CORO1B | 14 |
| GO:1901028 | regulation of mitochondrial outer membrane permeabilization involved in apoptotic signaling pathway | 6/294 | 34/9263 | 0.00061697 | 0.02540324 | 0.02282097 | YWHAZ/MPV17L/CHCHD10/HSPA1A/YWHAB/PPP3CC | 6 |
| GO:0090130 | tissue migration | 14/294 | 162/9263 | 0.00062695 | 0.0255177 | 0.02292379 | EMC10/PFN1/FOXP1/SH3BP1/TNFSF12/DOCK5/HDAC9/TMSB4X/JUN/GATA3/RGCC/ITGB1BP1/GIPC1/CORO1B | 14 |
| GO:0035148 | tube formation | 9/294 | 76/9263 | 0.00063713 | 0.02563715 | 0.0230311 | PFN1/CASP3/CFL1/LMO4/SEMA4C/GATA3/SKI/RARA/ITGB1BP1 | 9 |

| | | | | | | | | |
|---|---|---|---|---|---|---|---|---|
| GO:0043372 | positive regulation of CD4-positive, alpha-beta T cell differentiation | 5/294 | 23/9263 | 0.00065427 | 0.02603114 | 0.02338504 | NLRP3/IL18/IL23R/TNFSF4/RARA | 5 |
| GO:0002449 | lymphocyte mediated immunity | 15/294 | 182/9263 | 0.00066703 | 0.02624382 | 0.0235761 | NLRP3/CD81/IL18/HLA-B/CD28/LAMP1/BCL3/IL23R/HSPD1/TNFSF4/HLA-A/ARRB2/GATA3/EXOSC6/WAS | 15 |
| GO:0008637 | apoptotic mitochondrial changes | 10/294 | 93/9263 | 0.0007006 | 0.02726177 | 0.02449058 | YWHAZ/MPV17L/CHCHD10/HSPA1A/HSPD1/YWHAB/ARRB2/JUN/PPP3CC/IFI6 | 10 |
| GO:0035743 | CD4-positive, alpha-beta T cell cytokine production | 4/294 | 14/9263 | 0.00077356 | 0.02945354 | 0.02645955 | NLRP3/CD81/IL18/GATA3 | 4 |
| GO:0071526 | semaphorin-plexin signaling pathway | 4/294 | 14/9263 | 0.00077356 | 0.02945354 | 0.02645955 | SH3BP1/RAC1/ARHGDIA/SEMA4C | 4 |
| GO:0001667 | ameboidal-type cell migration | 16/294 | 205/9263 | 0.00080684 | 0.03039383 | 0.02730426 | EMC10/PFN1/FOXP1/SH3BP1/TNFSF12/DOCK5/CFL1/HDAC9/TMSB4X/JUN/SEMA4C/GATA3/RGCC/ITGB1BP1/GIPC1/CORO1B | 16 |
| GO:0030010 | establishment of cell polarity | 9/294 | 79/9263 | 0.00084717 | 0.03157699 | 0.02836715 | SH3BP1/CFL1/SPRY1/SLC9A3R1/MARK2/CDC42/GATA3/STK11/FAM89B | 9 |
| GO:0032496 | response to lipopolysaccharide | 17/294 | 228/9263 | 0.00093407 | 0.03413906 | 0.03066878 | JUNB/HSF1/CD6/FOXP1/CASP3/NFKBIB/NLRP3/HNRNPA0/IL18/IRAK3/ABCA1/PLCG2/IL23R/TNFSF4/JUN/PLSCR3/RARA | 17 |
| GO:0046637 | regulation of alpha-beta T cell differentiation | 7/294 | 50/9263 | 0.00093519 | 0.03413906 | 0.03066878 | NLRP3/IL18/RC3H1/IL23R/TNFSF4/GATA3/RARA | 7 |
| GO:2001233 | regulation of apoptotic signaling pathway | 20/294 | 291/9263 | 0.00094953 | 0.03430913 | 0.03082156 | YWHAZ/TRADD/BCL2L12/HSPA1B/GNAI2/DDX3X/MPV17L/MAL/TNFSF12/RRM2B/CHCHD10/HSPA1A/SLC9A3R1/TMC8/YWHAB/SKIL/A | 20 |

| | | | | | | | | |
|---|---|---|---|---|---|---|---|---|
| | | | | | | | RRB2/PPP3CC/CDKN2D/IFI6 | |
| GO:0043542 | endothelial cell migration | 11/294 | 114/9263 | 0.00096189 | 0.03440449 | 0.03090723 | EMC10/FOXP1/SH3BP1/TNFSF12/HDAC9/TMSB4X/GATA3/RGCC/ITGB1BP1/GIPC1/CORO1B | 11 |
| GO:0032233 | positive regulation of actin filament bundle assembly | 6/294 | 37/9263 | 0.00098464 | 0.03452097 | 0.03101187 | PFN1/SORBS3/RAC1/CDC42/RGCC/ITGB1BP1 | 6 |
| GO:0032720 | negative regulation of tumor necrosis factor production | 6/294 | 37/9263 | 0.00098464 | 0.03452097 | 0.03101187 | HSF1/IRAK3/BCL3/ARG2/ARRB2/RARA | 6 |
| GO:0051492 | regulation of stress fiber assembly | 7/294 | 51/9263 | 0.00105506 | 0.03662713 | 0.03290393 | PFN1/SORBS3/RAC1/CDC42/RGCC/ITGB1BP1/WAS | 7 |
| GO:0001838 | embryonic epithelial tube formation | 8/294 | 66/9263 | 0.00108608 | 0.0373378 | 0.03354236 | PFN1/CASP3/CFL1/LMO4/SEMA4C/GATA3/SKI/RARA | 8 |
| GO:0042692 | muscle cell differentiation | 13/294 | 153/9263 | 0.0011388 | 0.03848069 | 0.03456908 | CASP3/MORF4L2/CD81/FLT3LG/HDAC9/DNMT1/G6PD/PTBP1/CDC42/ARRB2/SEMA4C/SKI/RARA | 13 |
| GO:0002697 | regulation of immune effector process | 19/294 | 274/9263 | 0.00114105 | 0.03848069 | 0.03456908 | PCBP2/NLRP3/CD81/IL18/RC3H1/IRAK3/HLA-B/CD28/LAMP1/IL23R/HSPD1/TSPAN32/TNFSF4/HLA-A/ARRB2/GATA3/RARA/EXOSC6/WAS | 19 |
| GO:0032633 | interleukin-4 production | 5/294 | 26/9263 | 0.00118306 | 0.03919758 | 0.03521309 | NLRP3/CD28/TNFSF4/GATA3/RARA | 5 |
| GO:0001819 | positive regulation of cytokine production | 21/294 | 318/9263 | 0.00118445 | 0.03919758 | 0.03521309 | CD6/HSPA1B/IL17RA/NLRP3/CD81/IL18/DDX3X/LTB/CD28/HSPA1A/BCL3/TXK/PLCG2/IFNGR1/IL23R/HSPD1/TNFSF4/ADAM8/GATA3/RGCC/RARA | 21 |
| GO:0072175 | epithelial tube formation | 8/294 | 67/9263 | 0.00120007 | 0.03934686 | 0.0353472 | PFN1/CASP3/CFL1/LMO4/SEMA4C/GATA3/SKI/RARA | 8 |

| | | | | | | | | |
|---|---|---|---|---|---|---|---|---|
| GO:19 03556 | negative regulation of tumor necrosis factor superfamily cytokine production | 6/294 | 39/9263 | 0.0013 1033 | 0.042218 06 | 0.03792 654 | HSF1/IRAK3/BCL 3/ARG2/ARRB2/R ARA | 6 |
| GO:00 16331 | morphogene sis of embryonic epithelium | 8/294 | 68/9263 | 0.0013 2341 | 0.042218 06 | 0.03792 654 | PFN1/CASP3/CF L1/LMO4/SEMA4 C/GATA3/SKI/RA RA | 8 |
| GO:00 32649 | regulation of interferon-gamma production | 8/294 | 68/9263 | 0.0013 2341 | 0.042218 06 | 0.03792 654 | IL18/BCL3/TXK/IL 23R/HSPD1/TNF SF4/GATA3/RAR A | 8 |
| GO:00 02237 | response to molecule of bacterial origin | 17/294 | 236/926 3 | 0.0013 6499 | 0.043155 73 | 0.03876 889 | JUNB/HSF1/CD6/ FOXP1/CASP3/N FKBIB/NLRP3/HN RNPA0/IL18/IRAK 3/ABCA1/PLCG2/I L23R/TNFSF4/JU N/PLSCR3/RARA | 17 |
| GO:00 51347 | positive regulation of transferase activity | 25/294 | 412/926 3 | 0.0013 8916 | 0.043531 04 | 0.03910 605 | GADD45A/GADD 45G/CD81/IL18/U BE2S/DDX3X/MA PKAP1/TAOK3/D GKZ/JTB/MARK2/ RPS2/LMO4/PFK FB2/IL23R/LAT/T MSB4X/AXIN1/SE RINC5/ADAM8/C SK/RGCC/STK11/ ITGB1BP1/CALM 3 | 25 |
| GO:00 02369 | T cell cytokine production | 5/294 | 27/9263 | 0.0014 1474 | 0.043561 7 | 0.03913 359 | NLRP3/CD81/IL1 8/TNFSF4/GATA3 | 5 |
| GO:00 33120 | positive regulation of RNA splicing | 5/294 | 27/9263 | 0.0014 1474 | 0.043561 7 | 0.03913 359 | NUP98/HSPA1A/ SNRNP70/SF3B4/ U2AF2 | 5 |
| GO:00 30098 | lymphocyte differentiatio n | 18/294 | 258/926 3 | 0.0014 296 | 0.043639 67 | 0.03920 364 | FOXP1/NLRP3/IL 18/RC3H1/IRF2B P2/CD28/FLT3LG /HDAC9/BCL3/PL CG2/IL23R/TCF7/ TNFSF4/ZBTB7A/ ADAM8/GATA3/R ARA/STK11 | 18 |
| GO:00 42129 | regulation of T cell proliferation | 10/294 | 103/926 3 | 0.0015 4901 | 0.046823 46 | 0.04206 379 | CD6/RASAL3/CA SP3/CD81/IL18/R C3H1/CD28/IL23 R/TNFSF4/ARG2 | 10 |
| GO:00 10634 | positive regulation of epithelial cell migration | 9/294 | 86/9263 | 0.0015 6034 | 0.046823 46 | 0.04206 379 | EMC10/PFN1/FO XP1/DOCK5/HDA C9/TMSB4X/JUN/ GATA3/ITGB1BP 1 | 9 |
| GO:00 02699 | positive regulation of immune | 13/294 | 159/926 3 | 0.0016 179 | 0.048142 73 | 0.04324 895 | NLRP3/CD81/IL1 8/HLA-B/CD28/LAMP1/IL 23R/HSPD1/TNF | 13 |

| | | | | | | | | |
|---|---|---|---|---|---|---|---|---|
| | effector process | | | | | | SF4/HLA-A/GATA3/RARA/EXOSC6 | |
| GO:0110020 | regulation of actomyosin structure organization | 7/294 | 55/9263 | 0.00166 | 0.04898389 | 0.04400461 | PFN1/SORBS3/RAC1/CDC42/RGCC/ITGB1BP1/WAS | 7 |
| GO:0007163 | establishment or maintenance of cell polarity | 11/294 | 122/9263 | 0.00167725 | 0.04908393 | 0.04409448 | ANK1/SH3BP1/MAPKAP1/CFL1/SPRY1/SLC9A3R1/MARK2/CDC42/GATA3/STK11/FAM89B | 11 |
| GO:0032816 | positive regulation of natural killer cell activation | 4/294 | 17/9263 | 0.00170588 | 0.04951251 | 0.04447949 | IL18/FLT3LG/LAMP1/IL23R | 4 |

| Table S8: Antibodies Used in Flow Cytometry | | |
|---|---|---|
| **Targeting antigen** | **Company** | **Catalog/Clone** |
| Anti-Human BDCA1 | Biolegend | Cat# 354208 Clone: 201A (FITC) (1:200 dilution) |
| Anti-Human CD117 | Biolegend | Cat# 313206 Clone: 104D2 (APC) (1:200 dilution) |
| Anti-Human CD11c | Biolegend | Cat# 301604 Clone: 3.9 (FITC) (1:200 dilution) |
| Anti-Human CD123 | Biolegend | Cat# 306014 Clone: 6H6 (FITC) (1:200 dilution) |
| Anti-Human CD127 | Biolegend | Cat# 351320 Clone: A019D5 (PE/Cyanine7) (1:200 dilution) |
| Anti-Human CD14 | Biolegend | Cat# 325604 Clone: HCD14 (FITC) (1:200 dilution) |
| Anti-Human CD16 | Biolegend | Cat# 980104 Clone: 3G8 (APC) (1:400 diltuion) |
| Anti-Human CD19 | Biolegend | Cat# 302206 Clone: HIB19 (FITC) (1:200 dilution) |
| Anti-Human CD1a | Biolegend | Cat# 300104 Clone: HI149 (FITC) (1:200 dilution) |
| Anti-Human CD20 | Biolegend | Cat# 302304 Clone: 2H7a (FITC) (1:200 dilution) |
| Anti-Human CD22 | Biolegend | Cat# 363508 Clone: S-HCL-1 (FITC) (1:200 dilution) |
| Anti-Human CD3 | Biolegend | Cat# 317306 Clone: OKT3 (FITC) (1:200 dilution) |
| Anti-Human CD34 | Biolegend | Cat# 343504 Clone: 581 (FITC) (1:200 dilution) |
| Anti-Human CD4 | Biolegend | Cat# 317428 Clone: OKT4 (PerCP/Cyanine5.5) (1:200 dilution) |
| Anti-Human CD4 | Biolegend | Cat# 317408 Clone: OKT4 (FITC) (1:200 dilution) |
| Anti-Human CD45 | BD | Cat# 506178 Clone: 2D1 (APC/H7) (1:200 diltuion) |
| Anti-Human CD56 | Biolegend | Cat# 318306 Clone: HCD56 (PE) (1:200 dilution) |
| Anti-Human CD8 | Biolegend | Cat# 300924 Clone: HIT8a (PerCP/Cyanine 5.5) (1:200 dilution) |
| Anti-Human CRTH2 | Biolegend | Cat# 350116 Clone: BM16 (PerCP/Cyanine5.5) (1:200 dilution) |
| Anti-Human FcεR1α | Biolegend | Cat# 334608 Clone: AER-37 (FITC) (1:200 dilution) |
| Anti-Human TBX21 | ebioscience | Cat# 25-5825-82 Clone: ebio4B10 (PE/Cyanine7) (1:200 dilution) |
| Anti-Human TCRα/β | Biolegend | Cat# 306706 Clone: IP26 (FITC) (1:200 dilution) |
| Anti-Human TCRγ/δ | Biolegend | Cat# 331208 Clone: B1 (FITC) (1:200 dilution) |
| Anti-Human TCF7 | Cell Signaling | Cat# 37636s Clone: C63D9 (APC) (1:200 dilution) |
| Anti-Human IL-13 | Biolegend | Cat# 501908 Clone: JES10-5A2 (APC) (1:200 dilution) |
| Anti-Human AREG | ebioscience | Cat# 17-5370-42 Clone: AREG559 |
| mouse IgG1, k isotype control | Biolegend | Cat# 400112 Clone: MOPC-21 (PE) (1:200 dilution) |
| mouse IgG1, k isotype control | Biolegend | Cat# 400120 Clone: MOPC-21 (APC) (1:200 dilution) |
| Rabbit IgG, isotype control | Cell Signaling | Cat# 3452S (Alexa Fluor 647) (1:200 dilution) |
| Rat IgG1, k isotype control | Biolegend | Cat# 400412 Clone: RTK2071 (APC) (1:200 dilution) |

| **Table S9: Primers for bulk RNA-Seq** |
|---|
| **Barcoded primers for reverse transcription and final library amplification in bulk RNA-Seq** |
| Reverse transcription primers for 5 ILC sorted from blood (raw reads deposited in GSM5134071) |
| donor 1 ILCs: GCCGGTAATACGACTCACTATAGGGAGTTCTACAGTCCGACGATCNNNNNNGTTGCATTTTTTTTTTTTTTTTTTTTTTTTV |
| donor 2 ILCs: GCCGGTAATACGACTCACTATAGGGAGTTCTACAGTCCGACGATCNNNNNNACCATGTTTTTTTTTTTTTTTTTTTTTTTTV |
| donor 3 ILCs: GCCGGTAATACGACTCACTATAGGGAGTTCTACAGTCCGACGATCNNNNNNACTCGATTTTTTTTTTTTTTTTTTTTTTTTV |
| donor 4 ILCs: GCCGGTAATACGACTCACTATAGGGAGTTCTACAGTCCGACGATCNNNNNNACGTACTTTTTTTTTTTTTTTTTTTTTTTTV |
| donor 5 ILCs: GCCGGTAATACGACTCACTATAGGGAGTTCTACAGTCCGACGATCNNNNNNCTGTGATTTTTTTTTTTTTTTTTTTTTTTTV |
| Reverse transcription primers for 4 ILC samples sorted from blood (raw reads deposited in GSM5134072) |
| donor1 ILCs: GCCGGTAATACGACTCACTATAGGGAGTTCTACAGTCCGACGATCNNNNNNTCACCATTTTTTTTTTTTTTTTTTTTTTTTV |
| donor2 ILCs: GCCGGTAATACGACTCACTATAGGGAGTTCTACAGTCCGACGATCNNNNNNTCTGTCTTTTTTTTTTTTTTTTTTTTTTTTV |
| donor3 ILCs: GCCGGTAATACGACTCACTATAGGGAGTTCTACAGTCCGACGATCNNNNNNGTTGAGTTTTTTTTTTTTTTTTTTTTTTTTV |
| donor4 ILCs: GCCGGTAATACGACTCACTATAGGGAGTTCTACAGTCCGACGATCNNNNNNACAGACTTTTTTTTTTTTTTTTTTTTTTTTV |
| Final library amplification primers for all libraries: |
| RP1: AAT GAT ACG GCG ACC ACC GAG ATC TAC ACG TTC AGA GTT CTA CAG TCC GA |
| RPI1: CAA GCA GAA GAC GGC ATA CGA GAT CGT GAT GTG ACT GGA GTT CCT TGG CAC CCG AGA ATT CCA |
